# How local antibiotic use, carriage duration, resistance costs and international travel shape resistance frequency in E. coli in France

**DOI:** 10.64898/2026.04.16.26350860

**Authors:** Olivier Cotto, André Birgy, Mélanie Magnan, Stéphane Béchet, Stéphane Bonacorsi, Robert Cohen, Corinne Levy, Forough L. Nowrouzian, Olivier Tenaillon, François Blanquart

## Abstract

The worldwide rise in the prevalence of extended-spectrum beta-lactamase (ESBL) producing *Escherichia coli* is a major public health concern. In Europe, ESBL carriage frequency increased then stabilized at about 6-8 %. Past antibiotic use and travel in countries with high ESBL frequency, notably South-East Asia, have repeatedly been identified as risk factors of ESBL carriage. Yet, the relative contributions of these mechanisms to the observed maintenance of a stable low frequency of ESBL in Europe remains unknown. Here, we used comprehensive data on the risk factors for carriage of ESBL-producing *E. coli* in the French community, alongside detailed microbiological characterization of both resistant and overall *E. coli*, to develop a biologically plausible mathematical model of ESBL resistance spread in France. The model also includes several mechanisms previously showed to favor coexistence such as population structure, variability in carriage duration and within-host dynamics. The level of resistance in the community implies resistant strains transmit 14% less than sensitive (95% credible interval 0.6-38%), and are cleared at a +23% larger rate (0.9-62%). ESBL resistance is predicted to be strongly associated with factors prolonging residence in the gut. Both the rate of antibiotic treatment and transmission strongly impact the frequency of ESBL in the community. In contrast, travel has little impact on ESBL frequency. Whether reducing treatment or transmission is best to reduce resistance depends on community-specific parameters. Our study opens perspectives for the quantitative study of resistance evolution and argues for future work to improve the characterization of the duration of carriage of commensal bacterial strains.

## 1 Introduction

The worldwide rise in the prevalence of extended-spectrum beta-lactamase (ESBL) producing bacteria is a major health concern. These bacteria are resistant to many prescribed antibiotics, such as penicillins or third-generation cephalosporins and are frequently co-resistant to other antibiotic families like fluoroquinolones or trimethoprime/sulfamethoxazole, which leaves very few treatment options upon infection. Third-generation cephalosporin-resistant *Escherichia coli* caused an estimated 33,100 extra-deaths worldwide in 2021, making it one of the top ten deadliest pathogen-drug resistance combinations (Naghavi et al., 2024).

Epidemiological research has provided a comprehensive understanding of the risk factors and mechanisms underlying the spread of ESBL-resistant *E. coli*. While ESBL strains were initially mostly found in hospitals, from 2000 onward these strains have largely spread in the community as a whole (Pitout et al., 2005; Woerther et al., 2013). Frequencies of ESBL-producing *E. coli* are highly heterogeneous globally. In Europe, the ESBL carriage frequency in the community gently raised (Woerther et al., 2013; Bezabih et al., 2021) to stabilize from the 2010s at about 6-8 % (Blanquart, 2019; Bezabih et al., 2021; Castanheira et al., 2021; Pennings, 2025). In contrast, ESBL carriage increased to higher levels in other countries, in particular in South-East Asia, where studies commonly report a carriage of around 50%; carriage can reach values of 90% to 100% in some studies (reviewed in Woerther et al., 2013; Bezabih et al., 2021; Bui et al., 2015; Jacquier et al., 2023). Accordingly, in European communities, returning from South and Southeast Asia is an important risk factor for the carriage of ESBL-producing Enterobacterales (Tängdén et al., 2010; Kantele et al., 2015; Ruppé et al., 2015; Birgy et al., 2016; Karanika et al., 2016; Arcilla et al., 2017; Hu et al., 2020; Raffelsberger et al., 2023). Based on this observation, it has been argued that these travelers could potentially play an important role in shaping local levels of resistance (van der Bij and Pitout, 2012; Woerther et al., 2017; Olesen et al., 2020). Additionally, past antibiotic use was repeatedly identified as a risk factor for carriage of ESBL-producing E. coli (Birgy et al., 2016; Karanika et al., 2016; Hu et al., 2020; Raffelsberger et al., 2023), as is the case more generally for antibiotic-resistant bacteria (Chatterjee et al., 2018). In spite of this knowledge, the relative contributions of the different mechanisms in the observed maintenance of a stable low frequency of ESBL in Europe remain unknown (Emons et al., 2025).

Mechanistic mathematical models of resistance evolution are needed to help understand these contributions and eventually develop better public health policies to reduce resistances. Models are powerful tools to disentangle the role of interacting mechanisms, whether they are directly observed or not, on the dynamics of resistance, and assess the impact of management strategies. The impact of a reduction in antibiotic use (Lipsitch, 2001; Sundqvist et al., 2010; Blanquart et al., 2017; Emons et al., 2025) and the impact of a reduction in transmission (Davies et al., 2019; Jacopin et al., 2020) on resistance frequency levels both depend on modelling assumptions. For example, in the case of the impact of transmission, transmission can favor the resistant strain by enabling it to transmit to empty treated hosts. But it can also favor the sensitive strain by allowing it to supercolonize and displace the resistant one. The end result of these two processes vary depending on the assumptions on co-colonization. Thus, mechanistic models with robust assumptions are needed to anticipate both future evolution and impacts of interventions.

Intriguingly, there are few mechanistic mathematical models of ESBL resistance evolution in *E. coli* (Rahbé et al., 2024). Morevoer, most of these models suffer either from assumptions that are not supported by biological data, and/or from predicted behaviors that are not consistent with the observed resistance dynamics. One central set of assumptions regards the description of competition between strains. Competition between strains profoundly shapes resistance dynamics (Lipsitch et al., 2009; Blanquart, 2019; Davies et al., 2019). It is biologically intuitive that strains of the same species that only differ in their resistance status should occupy the same niche in their host, and thus compete for nutrient or space. This has been recently shown for *E. coli* in multiple datasets (Morel-Journel et al., 2025): *E. coli* strains are cleared faster when several strains compete simultaneously in the host. Moreover, ESBL resistant strains are most often coexisting with sensitive strains at low density (Ruppé et al., 2013; de Lastours et al., 2016). In spite of these observations, half of existing models of antibiotic-resistance in *Enterobacterales* describe only the resistant strain (Rahbé et al., 2024) which questions the plausibility and applicability of their predictions. Studies that correctly include the competition between sensitive and resistant strains often do not reproduce the observed coexistence of these strains–the so-called “coexistence problem” (Lipsitch et al., 2009). Instead, models predict that either the sensitive or resistant strain replaces the other in most conditions (this is the case for models in Sundqvist et al. 2010; MacFadden et al. 2019; Godijk et al. 2022). A notable exception is the work of Kachalov et al. (2021) on *K. pneumoniae*; they modelled the dynamics of ESBL resistance in hospitals and the community and reproduced the frequency of resistance across eleven European countries.

The coexistence of resistant and sensitive strains at the community scale, as observed in *E. coli*, poses a more general theoretical problem (Blanquart, 2019). Several theoretical solutions have been proposed (Colijn et al., 2010), including host population structure (Cobey et al., 2017; Blanquart et al., 2018), variability in the duration of carriage of bacterial strains (Lehtinen et al., 2017), slow dynamics of exclusion of the resistant strains by the sensitive strain within hosts (Davies et al., 2019). However, these theoretical models are either not precisely parameterized with data, or the parameterization of these models with existing data fails to fully explain coexistence. This failure can be explained in two ways. A first possibility is that yet unknown factors play a major role in the stabilization of resistance frequencies. A second possibility is that a combination of the aforementioned solutions together would explain coexistence. Parameterizing complex models is difficult and requires sufficiently detailed data on a system. This might have hindered progress towards a satisfactory solution combining several mechanisms.

In this study, we use comprehensive data on the risk factors for carriage of ESBL-producing *E. coli* in the French community, alongside detailed microbiological characterization of both resistant and overall *E. coli*, to develop a biologically plausible mathematical model of ESBL resistance spread in France combining several mechanisms stabilizing coexistence. Earlier analyses identified travel to South-East Asia and prior use of third-generation cephalosporins as key risk factors for ESBL *E. coli* carriage, consistent with other studies (Birgy et al., 2016). Here, we use this dataset extended to three more years, and additionally quantify the total Enterobacterales density and the density of ESBL-producing Enterobacterales in a subset of samples. This allows us to precisely model competition by inferring the prevalence of different colonization states: resistant-only, sensitive-only, and co-colonization. We model both the impact of travel to countries with a high prevalence of ESBL, and the impact of treatment clearing sensitive strains, with the aim to reproduce the inferred risk factors for ESBL carriage. We also model the known variability in carriage duration across *E. coli* strains (Östblom et al., 2011; Martinson et al., 2019; Morel-Journel et al., 2023), and plausible within-host dynamics of resistant and sensitive strains (Cotto et al., 2023). Thus, all in all, our study includes the main theoretical explanations for coexistence: population structure, variability in the duration of carriage, and within-host dynamics. We model and infer in a Bayesian framework multiple fitness costs associated with epidemiological traits such as transmission and clearance of resistant strains. We verify the goodness-of-fit of our model and finally predict how changes in the rate of travel, the transmission rate and antibiotic use influence resistance levels in the community.

## 2 Methods

### 2.1 Data and model structure

Our main data set is an epidemiological study of ESBL-producing *E. coli* in rectal samples from *N* = 3443 French children aged 6-24 months from 2010 to 2018. Rectal samples were provided during visits to pediatricians on a voluntary basis. Sampling was associated with a questionnaire providing information on behaviors prior to sampling, including travel and antibiotic treatment. Children undergoing antibiotic treatment within 7 days before sampling were excluded from the survey. All samples were screened for the ESBL-producing Enterobacterales for a total of *N* = 257 resistant strains found. The details of the survey can be found in Birgy et al. (2016).

We focused on children to benefit from an established network of pediatricians for the collection of rectal samples. The literature on ESBL resistance in Europe suggests that the prevalence of this resistance does not differ much between age classes. Indeed, mean estimates of ESBL prevalence range from 3.5 to 7.6% in preschool children (Van den Bunt et al., 2016; Birgy et al., 2016), similar to those estimated in workers and retired age classes (6-8.6 %, Nicolas-Chanoine et al. 2013; Reuland et al. 2016).

#### 2.1.1 Risk factors associated with ESBL carriage

A previous statistical analysis of a subset of the data (from 2010 to 2015, Birgy et al. 2016) revealed that children who have traveled to South-East Asia (SEA) or who have received betalactam antibiotics within a three-months period preceding the sampling are more likely to carry ESBL-producing strains. We replicated the analyses of (Birgy et al., 2016) to our extended data set (2010-2018) and found similar results (Supp.Mat. S1).

Following the result of the statistical analysis (Birgy et al. 2016 and Supp.Mat. S1) and previous results on risk factors for ESBL carriage, we modeled five compartments to describe antibiotic treatment and travel to SEA (Figure 1A). The first compartment, “untreated” *U* counts individuals who neither traveled nor used antibiotics. Individuals receive antibiotic treatment with rate *τ* and subsequently enter compartment “treated” *T*. Antibiotic treatment includes all types of beta-lactams (see Supp. Mat. S1). Treatment stops at rate *ω* and individuals enter compartment “three months post-treatment” *T*_3_ where they remain on average three months (they leave *T*_3_ at rate *ω*_3_ = 1/3 month^−1^). Travel to SEA was modeled similarly to antibiotic treatment. Individuals travel to SEA at rate *ν*. While traveling, individuals are in compartment “South-East Asia” *SEA* where they remain for a period measured by *ψ*^−1^ where *ψ* is the rate of return from travel. After travel, these individuals remain in compartment “three months post South-East Asia” *SEA*_3_ for three months (they leave *SEA*_3_ to compartment *U* at rate *ψ*_3_ = 1/3 month^−1^). For mathematical convenience, we assumed a standard exponential distribution residency time in *T*_3_ and *SEA*_3_. For simplicity, individuals can be either in compartment *SEA* or *T*, such that we did not consider individuals that both traveled and used antibiotics.

**Figure 1:**
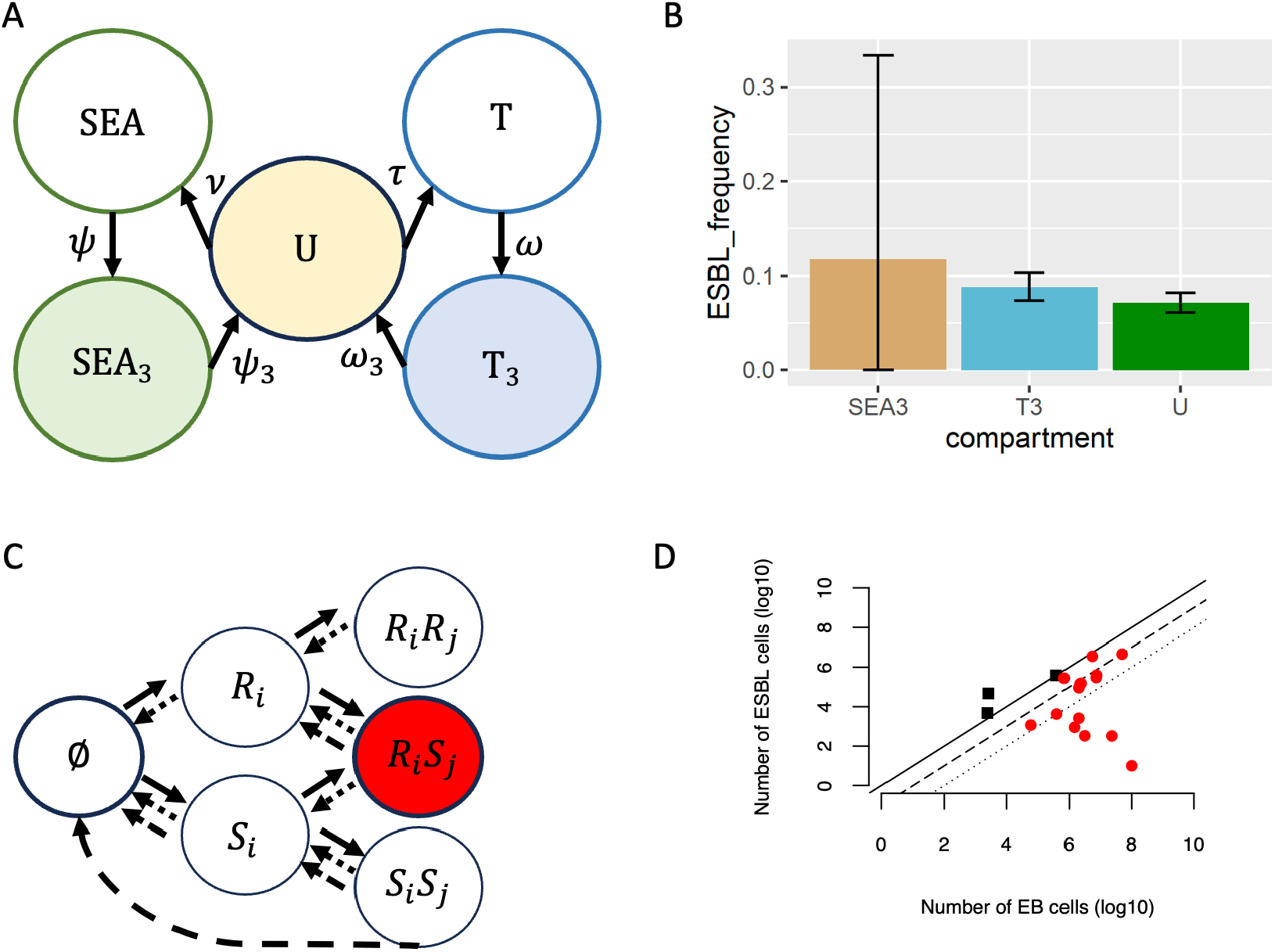
Data-driven epidemiological model. A) compartments describing host behaviors and sampling. *U* : individuals who did not travel nor used antibiotics, *T* : individuals under antibiotic treatment, *SEA*: individuals traveling to South-East-Asia. the subscript 3 indicates that the corresponding behavior occurred within 3 months before sampling. Only the compartments U, SEA3 and T3 are observed (sampled). B) ESBL frequencies in the three observed host behaviors. C) Structure of the multiple-carriage model. Hosts can be colonized by up to two strains that belong to type *C* or *P* (*i, j* ∈ {*C, P*}). Estimate of Enterobacterales density in 301 samples allowed to estimate the frequency of uncolonized (thick-open) and mixed-carriage hosts (thick-red). Solid, dashed and dotted arrows represent colonization, natural clearance and antibiotic-driven clearance, respectively. D) Density of ESBL-carrying Enterobacterales as a function of the total density of Enterobacterales. Dark square: all Enterobacterales in the sample carried ESBL (R carriage hosts); red-filled dots: mixed-carriage (RS) hosts. Solid, dashed and dotted lines: fraction of ESBL-carrying strains equals 100%, 10% and 1% of the total.

We used the frequencies of ESBL carriers in the (observed) compartments *U, SEA*_3_ and *T*_3_ for parameter inference (Figure 1).

#### 2.1.2 Multiple carriage

We developed a structurally neutral, mixed-carriage model (sensu Davies et al., 2019) to describe the epidemiological dynamics in the system of compartments described above (Figure 1C). Individuals can be colonized by up to two strains. We assumed that strains can be either sensitive (S) or resistant (R) to beta-lactams.

In the initial model, we considered two strain types, R and S. After inference of the parameters (see below), the model was able to reproduce the observed equilibrium, but coexistence between R and S strains occurred for a narrow range of parameters and the dynamics of equilibration was much slower than the observed one (presented in Supp. Mat. S7). To improve the model, we introduced further genetic structuring by modelling a locus underlying within-host persistence, as theory showed that variability in duration of carriage across different bacterial genotypes can be a factor maintaining coexistence (Lehtinen et al., 2017). At this locus, we considered colonizer (C) and persistent (P) types. Including this genetic structuring in the model was further supported by recent findings in *E. coli* showing that strain types locate along a colonization-persistence trade-off (Morel-Journel et al., 2025). Further, ESBL-carriage appear to be more frequent in phylogroups B2, D, F and particularly in sequence type ST131 (Birgy et al., 2016), all associated with long carriage time (Östblom et al., 2011; Johnson et al., 2022; Martinson et al., 2019). Overall, we considered four types of strains, which are {*S*_*C*_, *S*_*P*_, *R*_*C*_, *R*_*P*_}, where the capital letter and index characterize resistance to beta-lactams and gut persistence, respectively. It follows that individuals can be either uncolonized (1 possibility), single colonized by one of the four strains (4 possibilities), or co-colonized (fully sensitive *S*_*k*_*S*_*l*_, fully resistant *R*_*k*_*R*_*l*_, or mixed *R*_*k*_*S*_*l*_, *k* and *l* in {*C, P*}, 10 possibilities), which all together correspond to 15 possible colonization statuses. Resistance corresponds to ESBL-producing strains

To inform the description of co-colonizations in our model, we inferred from our data what fraction of hosts were uncolonized by Enterobacterales, colonized only by ESBL strains, or by ESBL together with sensitive strains (Fig. 1C). To do so, among the *N* = 3443 samples, we randomly selected 301 samples to measure the total density of Enterobacterales (EB) together with the density of ESBL-carrying EB (Supp.Mat.S2). Samples where no EB were found were considered uncolonized. Samples containing both ESBL and sensitive strains were considered as mixed-carriage individuals (Fig. 1D). Lastly, samples containing only ESBL strains were considered as R individuals, with either only one or two R strains. The frequencies of uncolonized and mixed-carriage hosts were used for parameter inference.

Previous analyses showed that clearance of resistant strains occurs as an abrupt process following a potentially long period of carriage (Cotto et al., 2023). Change of within-host densities of resistant and sensitive strains was not observable at the time scale of that study. Such dynamics are a key determinant of the between-host epidemiological dynamics (Davies et al., 2019). In accordance with our results, we modeled competition between resistant and sensitive strains as a clearance-recolonization process (Figure 1C). Strains are occasionally cleared from the gut of their hosts, thus opening opportunities for re-colonization. Strains compete to colonize new hosts and reside in them (competition at the between-host scale).

All in all, the epidemiological dynamics is described by a system of ordinary differential equations (ODE) defining the changes in the frequencies *X*_*j,i*_ of the 75 types of hosts, where the index *j* is a pair in the set {∅, *S*_*C*_, *S*_*P*_, *R*_*C*_, *R*_*P*_} indicating the colonization status as defined above (15 possible pairs), and the index *i* corresponds to one of the five epidemiological compartments {*U, T, T*_3_, *SEA, SEA*_3_}. The full system of equations is available as an online supplementary material. The model was simulated until equilibrium was reached. Comparisons between the frequencies predicted by the model at equilibrium and the five statistics in the data (frequencies of ESBL in *U, SEA*_3_, *T*_3_ and frequencies of uncolonized and mixed-carriage hosts) in a likelihood framework were used to infer the parameters of interest (see below).

### 2.2 Parameters

#### 2.2.1 Models for the transmission and clearance rates

##### Transmission and clearance rates in relation to the colonization status

Transmission and clearance rates in principle depend both on the strain and the type of host. For inference, we reduced the number of parameters based on biologically plausible assumptions (Table 1). First, we modeled the transmission rate of a given strain as depending only on whether the recipient host is already colonized. We note 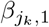 and 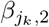, the colonization by strain *j*_*k*_ of uncolonized and single-colonized hosts, respectively.

**Table 1:**
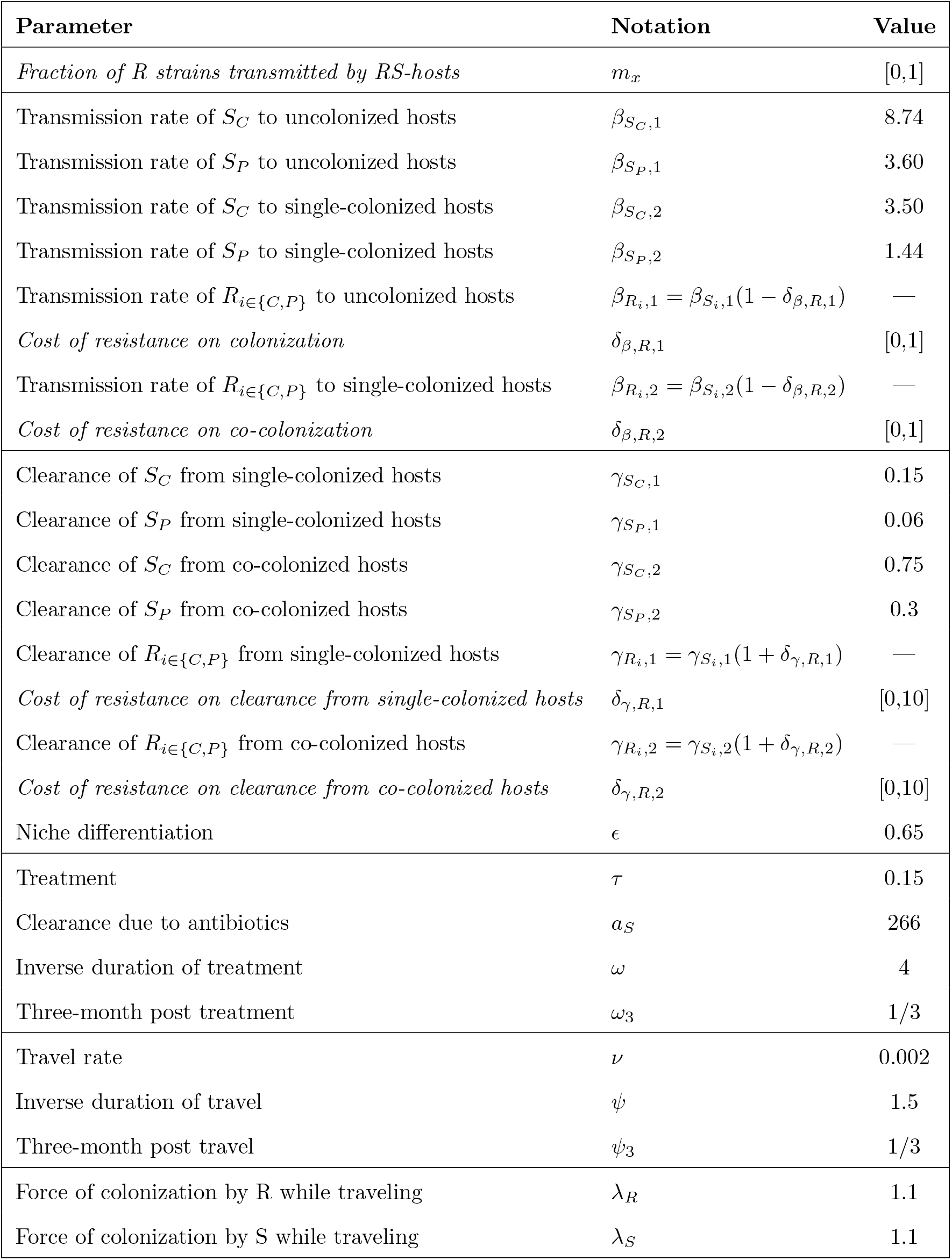
Parameters of the model. Parameters to be inferred (in the set *P*_*R*_) are in italic. In the column “Value”, bracket intervals denote the ranges investigated. We used uniform prior within these ranges for the parameters in *P*_*R*_. Hyphens indicate that the value of the parameters is defined by the value of the *δ*’s. Rate parameters are in units of months^−1^. See also Supp.Mat. S3

| Parameter | Notation | Value |
| --- | --- | --- |
| *Fraction of R strains transmitted by RS-hosts* | $m_x$ | [0,1] |
| Transmission rate of $S_C$ to uncolonized hosts | $\beta_{S_C,1}$ | 8.74 |
| Transmission rate of $S_P$ to uncolonized hosts | $\beta_{S_P,1}$ | 3.60 |
| Transmission rate of $S_C$ to single-colonized hosts | $\beta_{S_C,2}$ | 3.50 |
| Transmission rate of $S_P$ to single-colonized hosts | $\beta_{S_P,2}$ | 1.44 |
| Transmission rate of $R_{i \in \{C,P\}}$ to uncolonized hosts | $\beta_{R_i,1} = \beta_{S_i,1}(1 - \delta_{\beta,R,1})$ | — |
| *Cost of resistance on colonization* | $\delta_{\beta,R,1}$ | [0,1] |
| Transmission rate of $R_{i \in \{C,P\}}$ to single-colonized hosts | $\beta_{R_i,2} = \beta_{S_i,2}(1 - \delta_{\beta,R,2})$ | — |
| *Cost of resistance on co-colonization* | $\delta_{\beta,R,2}$ | [0,1] |
| Clearance of $S_C$ from single-colonized hosts | $\gamma_{S_C,1}$ | 0.15 |
| Clearance of $S_P$ from single-colonized hosts | $\gamma_{S_P,1}$ | 0.06 |
| Clearance of $S_C$ from co-colonized hosts | $\gamma_{S_C,2}$ | 0.75 |
| Clearance of $S_P$ from co-colonized hosts | $\gamma_{S_P,2}$ | 0.3 |
| Clearance of $R_{i \in \{C,P\}}$ from single-colonized hosts | $\gamma_{R_i,1} = \gamma_{S_i,1}(1 + \delta_{\gamma,R,1})$ | — |
| *Cost of resistance on clearance from single-colonized hosts* | $\delta_{\gamma,R,1}$ | [0,10] |
| Clearance of $R_{i \in \{C,P\}}$ from co-colonized hosts | $\gamma_{R_i,2} = \gamma_{S_i,2}(1 + \delta_{\gamma,R,2})$ | — |
| *Cost of resistance on clearance from co-colonized hosts* | $\delta_{\gamma,R,2}$ | [0,10] |
| Niche differentiation | $\epsilon$ | 0.65 |
| Treatment | $\tau$ | 0.15 |
| Clearance due to antibiotics | $a_S$ | 266 |
| Inverse duration of treatment | $\omega$ | 4 |
| Three-month post treatment | $\omega_3$ | 1/3 |
| Travel rate | $\nu$ | 0.002 |
| Inverse duration of travel | $\psi$ | 1.5 |
| Three-month post travel | $\psi_3$ | 1/3 |
| Force of colonization by R while traveling | $\lambda_R$ | 1.1 |
| Force of colonization by S while traveling | $\lambda_S$ | 1.1 |

We also simplified natural clearance rates. The clearance rate of a given strain depends on whether the host is single- or co-colonized. The clearance rate from a host single-colonized by strain *j*_*k*_ is 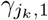. For co-colonized hosts, we assumed niche differentiation: clearance of one strain is slower when it co-occurs in the host with a strain of the other type (C or P). Niche differentiation between colonizer and persistent strains is measured by parameter *ϵ* ∈ [0, 1]. The clearance rate of strain *j*_*k*_ from a host co-colonized by strains *j*_*k*_ and *i*_*l*_ (with *k, l* ∈ *C, P*) is 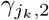 if *l* = *k*, and 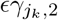 otherwise (i.e., clearance is reduced by a factor *ϵ* if the co-occurring strains are of a different type). We further assumed that clearance rates are the same in all epidemiological compartments.

We did not explicitly model the epidemiological dynamics abroad. Instead, we modeled a constant force of colonization while traveling, equal to *λ*_*y*_, where *y* is the colonizing strain.

##### Transmission rate from co-colonized hosts

We modeled hosts as two “spaces”, each of which can be colonized by a single strain. This assumption could result, for example, from the fine-scale spatial structure of the niche occupied by *E. coli* in the gut (e.g. Pereira and Berry 2017). We assumed that co-colonized hosts transmit twice as much as single-colonized hosts, such that the transmission rate from {*y, y*}-hosts is 2*X*_*y,y*_*β*_*y,i*_ (*i* ∈ 1, 2). We performed an analysis of sensitivity to this assumption and found that it did not affect our main conclusions (Supp.Mat.S6).

For simplicity, we neglected the probability of co-transmission by co-colonized hosts. Further, we modeled that mixed-carriage hosts carrying both a resistant (R) and a sensitive (S) strain can shed these strains at different rates, scaled by the parameter *m*_*x*_. Precisely, {*R*_*k*_, *S*_*l*_}-hosts transmit R strains at rate 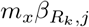 and S strain at rate 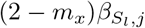, where *j* ∈ {1, 2} and *k, l* ∈ {*C, P*}. Given that we expect *R* strains to occur at low frequency relative to *S* strains (Ruppé et al., 2013; de Lastours et al., 2016), we assume that *m*_*x*_ ∈ [0, 1]. The value *m*_*x*_ = 1 corresponds to R strains in mixed-carriage transmitting as much as if they were alone in the host.

##### Transmission and clearance rates of resistant strains

The epidemiological parameters of resistant strains are defined relative to those of sensitive strains through the cost parameters *δ*s acting on the transmission and clearance rates. The costs of resistance did not depend on whether it was associated with a persistent (P) or colonizer (C) type. Given 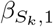 and 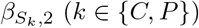, we distinguish the cost of resistance on colonization and super-colonization as

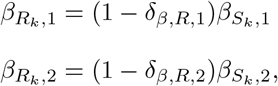

where *δ*_*β,R*,1_ and *δ*_*β,R*,2_ are both included in [0, 1] and measure the cost of resistance on colonization and super-colonization, respectively. Similarly, we assumed that resistance can reduce persistence through an increased clearance rate that depends on whether the host is single- or co-colonized.

We modeled the clearance rates of resistant strains compared to that of sensitive strains as

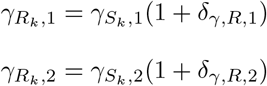

where *δ*_*γ,R*,1_ and *δ*_*γ,R*,2_ are the costs of resistance included in [0, ∞].

#### 2.2.2 Parameter estimation

Overall, we assume that the costs of resistance can occur on transmission and clearance and that they can depend on the colonization status of the hosts. We infered the costs of resistance that are measured by the set of parameters :

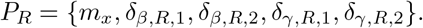

The other parameters were directly obtained from the focal data or from data in similar communities, as broadly described below. The details of the parameter estimation are provided in Supplementary Material S3. Table 1 recapitulates the model parameters.

##### Rates of travel to SEA and antibiotic use

These rates were directly estimated from the mean per-year fraction of children traveling to SEA and having used antibiotics [including Cefpodoxime N=144 (11%), Amoxicillin N=681(54%), Amoxicillin and clavulanic acid N=370 (29%), others N=75 (6%)] in the three months before sampling in our epidemiological dataset.

##### Force of infection in South-East Asia

We assumed a fixed force of new colonization by sensitive and resistant strains in South-East Asia, denoted *λ*_*S*_ and *λ*_*R*_, respectively. The value of *λ*_*R*_ was chosen such that about 50% of travelers would be colonized by R-strains during a 20-days travel (see Supp.Mat.S3). Without information on the colonization rate by S-strains during travel, we assumed *λ*_*S*_ = *λ*_*R*_.

##### Epidemiological parameters of sensitive strains

Our focal data set did not allow inference of the colonization and clearance rates of sensitive strains. To do so, we used data on the turnover of *E. coli* strains in Swedish children from birth to 12 months old from Östblom et al. (2011). We assumed this Swedish children community had similar characteristics to our focal one. The analysis of these data is provided in Supplementary Material S3. These data allowed to estimate the clearance rates of sensitive strains from single- and from co-colonized hosts 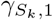 and 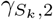 for both the colonizer and persistent types (*k* ∈ {*C, P*}), and the niche differentiation parameter *ϵ* (Table 1). The persistent (P) and colonizer (C) types were identified as belonging and not belonging to the phylogroup B2, respectively, consistent with the results of Östblom et al. (2011). We then computed the transmission rates such that strains *S*_*C*_ and *S*_*P*_ had the same *R*_0_ (fitness) and could therefore readily coexist in the community.

##### Inference of the epidemiological cost parameters of resistant strains

We used a Bayesian approach to infer the posterior distribution of the cost parameters in *P*_*R*_. We formulated the likelihood with five independent components, using two pieces of information derived from our focal dataset. For each component, the likelihood of the observed numbers (*n*_*R,i*_) is their probabilities under the law *B*(*N*_*i*_, *p*_*i*_) with *N*_*i*_ the total number of observations in component *i* and *p*_*i*_ the corresponding frequency predicted by the model. First, our epidemiological data inform on the frequency of resistance in compartments *U* (*n*_*R,U*_ = 105, *N*_*U*_ = 1632), 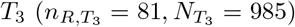 and *SEA*_3_ 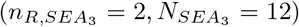. Note that we did not consider individuals both returned from travel and who used antibiotics. We additionally measured the density of Enterobacterales and ESBL-producing *E. coli* on a sub-sample of the data (*N* = 301, Supp.Mat S2) to inform on the overall frequency of mixed-carriage hosts *X*_*RS*_ (*n*_*RS*_ = 15) and uncolonized hosts *X*_∅_ (*n*_∅_ = 5).

Posterior distributions of the parameters were estimated using a Bayesian approach with flat priors (see Table 1). The posterior distributions were sampled by MCMC using a Metropolis-Hastings algorithm with adaptive sampling (Supp.Mat.S4). We ran five independent MCMC chains of 250000 iterations to check convergence to the same posterior distributions. For each chain, the initial conditions were drawn randomly within the prior distribution (Table 1). The effective sample size and the convergence of each chain were verified using the Gelman-Rubin diagnostic implemented in the R-package coda (Plummer et al. 2006). Statistics on the MCMC chains are provided in Supplementary Material S5.

## 3 Results

We first present the epidemiological data, before turning to parameter inference with the model and its sensitivity analysis. The frequency of resistance in the community was defined as the frequency of hosts who carry at least one resistant strain.

### 3.1 ESBL carriage

Over 2010-2018, ESBL were found in 257 of the 3443 assessed children (7.5%, 95% CI=6.7-8.4). There was no trend in the change in ESBL during the study period (black on Fig. 2), as the increase in ESBL frequency in the community occurred before that period (blue on Fig.2). The ESBL frequency was particularly stable over the period 2015-2019. Among the assessed children, 0.6 % had travelled to SEA and 37% had received antibiotics within seven to three months before enrollment. Further details are provided in tables S1 and S2.

**Figure 2:**
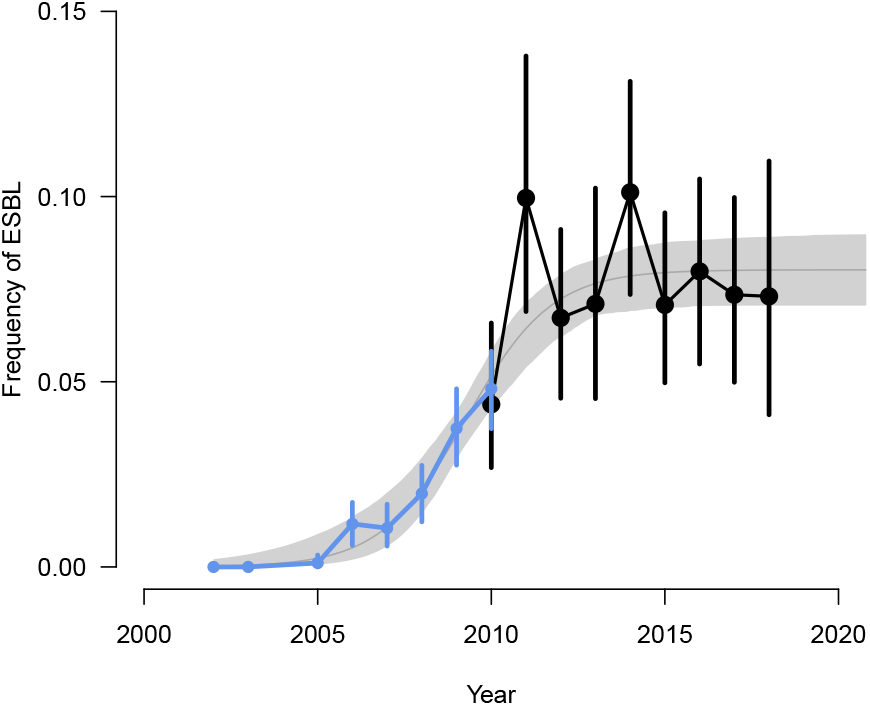
ESBL carriage as a function of time. Black: ESBL frequency in the community, measured from French children during the period 2010-2018 (focal data set). Blue: ESBL frequency in outpatients in data from the European Center for Diseases prevention and Control (ECDC), available for outpatients only over the period 2002-2010. Data are mean percentages with 95% confidence intervals. The thin gray line is a logistic model with a plateau fitted to these data with least squares, with weights proportional to the number of isolates at each timepoint. The gray envelope shows the 95% confidence intervals.

### 3.2 Inference of the single and mixed carriage of EBSL

To inform the mixed-carriage model, we inferred the total number of Enterobacterales (mostly and hereafter *E. coli*) and the number of those carrying ESBL in a subset of 301 samples (details in Supp. Mat. S2). We found that, in most cases, ESBL and non-ESBL *E. coli* co-occurred in the samples (Fig. 1D). Indeed, in most (*N* = 15) of the samples containing ESBL-carrying *E. coli* (*N* = 18, see Tab. S3), the estimated number of ESBL-carrying *E. coli* was lower than the estimated total number of *E. coli*. In these samples, ESBL-carrying *E. coli* represented on average 4.8 % of the total number of *E. coli*. In the remaining ESBL-carrying samples (*N* = 3), the ESBL-carrying E. coli formed the majority of E. coli (Tab. S3). Lastly, we found five samples that were apparently not colonized by any *E. coli*.

### 3.3 Inference of the costs of resistance

#### Goodness of fit

Overall, after inference, the model was able to reproduce the frequencies of the different types of host colonization observed in the focal data set. The distribution of the host-type frequencies predicted by the model after sampling the posterior distribution falls mainly within the 95% confidence interval of the observed frequencies (Fig.S10).

#### Costs of resistance on transmission and clearance

We found that resistance is costly: R-strains transmit less efficiently and are cleared faster than S-strains (Fig. 3A-D). The costs of resistance depended on the colonization status of the hosts. Our model was able to infer relatively precise estimates of the costs of resistance on colonization of already colonized hosts and on persistence in co-colonized hosts (i.e., additional clearance). These costs must be low to explain the data. Precisely, we inferred the cost on the ability to colonize already-colonized hosts (*δ*_*β,R*,2_) at 0.14 (median = 0.12, 95% CI = 0.006-0.38, Fig. 3B); and the cost on the persistence ability in co-colonized hosts (*δ*_*γR*,2_) at 0.23 (median = 0.20, 95% CI = 0.009-0.62, Fig. 3D). In other words, resistant strains colonize occupied hosts at a rate reduced by −14%, and are cleared +23% faster when in competition with other strains. These two costs are the most precisely inferred, because they concern the most common ecological contexts in which resistant strains are encountered.

**Figure 3:**
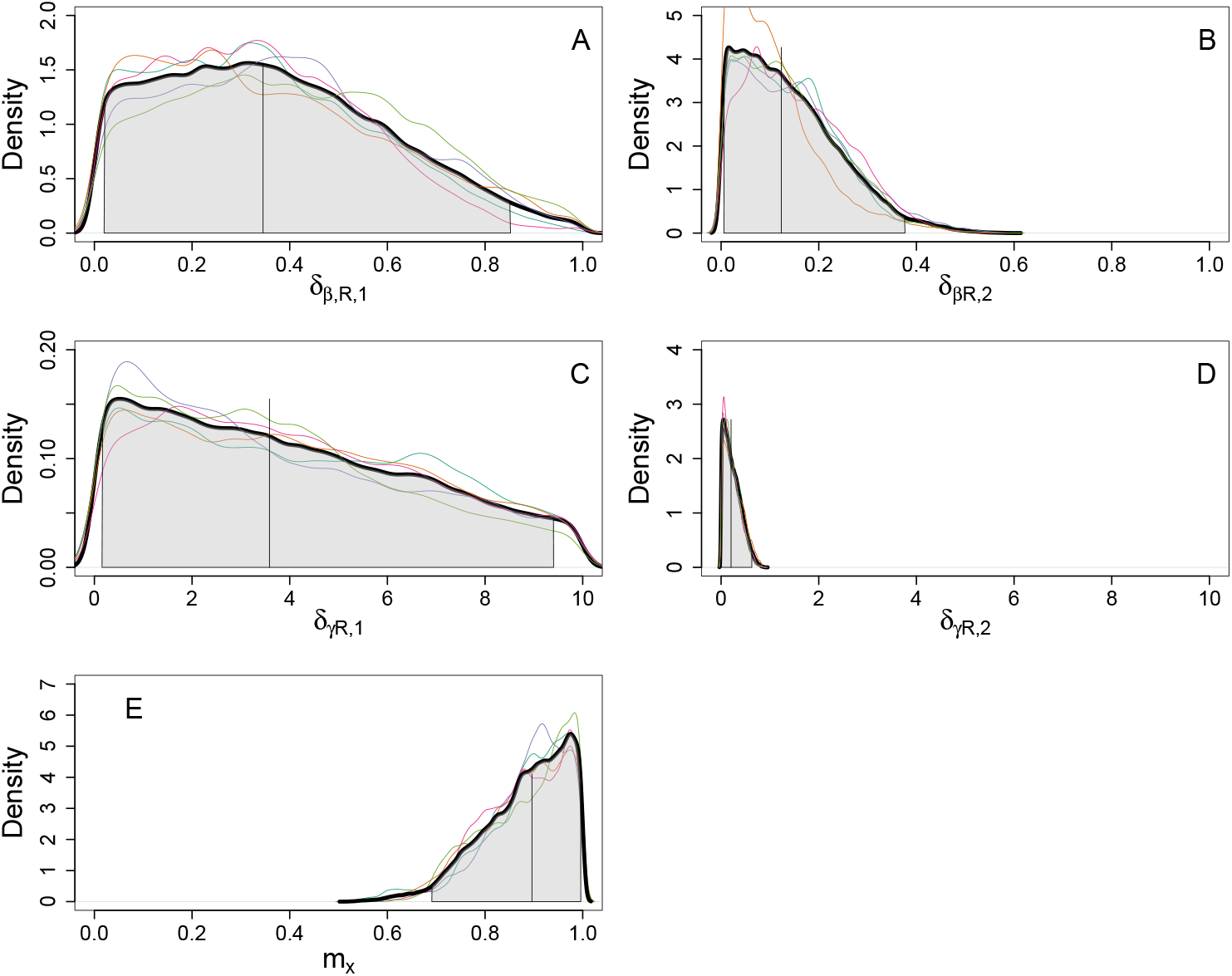
Posterior distribution of the five estimated parameters. For all panels : Colored curves : posterior distribution for each of the five MCMC chains. Black curve: all chains pooled. Vertical line: Mean of the distribution. Shaded area: 95% CI. A and B, costs of resistance on colonization of uncolonized *δ*_*β,R*,1_ and single-colonized hosts *δ*_*β,R*,2_, respectively. C and D, additional clearance of resistant strains from single colonized *δ*_*γ,R*,1_ and co-colonized hosts *δ*_*γ,R*,2_, respectively. E: *m*_*x*_. costs of resistance on colonization and clearance. The prior distributions of all parameters are uniform on the intervals [0,1], [0,1], [0,10], [0,10], and [0,1], respectively.

In contrast, the posterior distributions of the costs on transmission to uncolonized hosts and clearance from single-colonized hosts were much wider. The 95% CI of the posterior distribution span most of the prior distribution for the parameters *δ*_*β,R*,1_ and *δ*_*γ,R*,1_ (shadowed area in Fig. 3A and C). A wide range of resistance costs on colonization of uncolonized hosts and persistence in single-colonized hosts are compatible with the data.

#### Contributions of mixed-carriage to transmission

We found that the transmission rate of a R-strain is similar when it is in mixed carriage with a S-strain and when it is alone. Indeed, we estimated that the mean transmission rate of a R-strain while in mixed carriage (*m*_*x*_ parameter) is 88% (median 90, 95% CI = 68-99) of that when it is alone (Fig. 3E). This is consistent with the analysis of the count data showing that the density of ESBL-carrying *E. coli* is similar in hosts colonized only by these strains and in those carrying both ESBL and non-ESBL *E. coli* (Fig. 1D).

Overall, given the inferred mean values of the different fitness costs of resistance, our model predicts that after stopping travel and antibiotic treatment, it would take approximately 30 (resp. 60) months for the frequency of antibiotic resistance in the community to decline below 1% (resp. 1/1000).

### 3.4 Predictions of the model

Next, we investigated the dynamical consequences of changes in key parameters of the model on the frequency of resistance in the community. The reference parameter values were those of Table 1 completed by the means of the posterior distributions for the inferred parameters (Fig. 3). Resistance frequency was mainly driven by the treatment rate and the transmission rate of *E. coli* (Fig. 4A and B), but not by travel (Fig. 4A and C).

**Figure 4:**
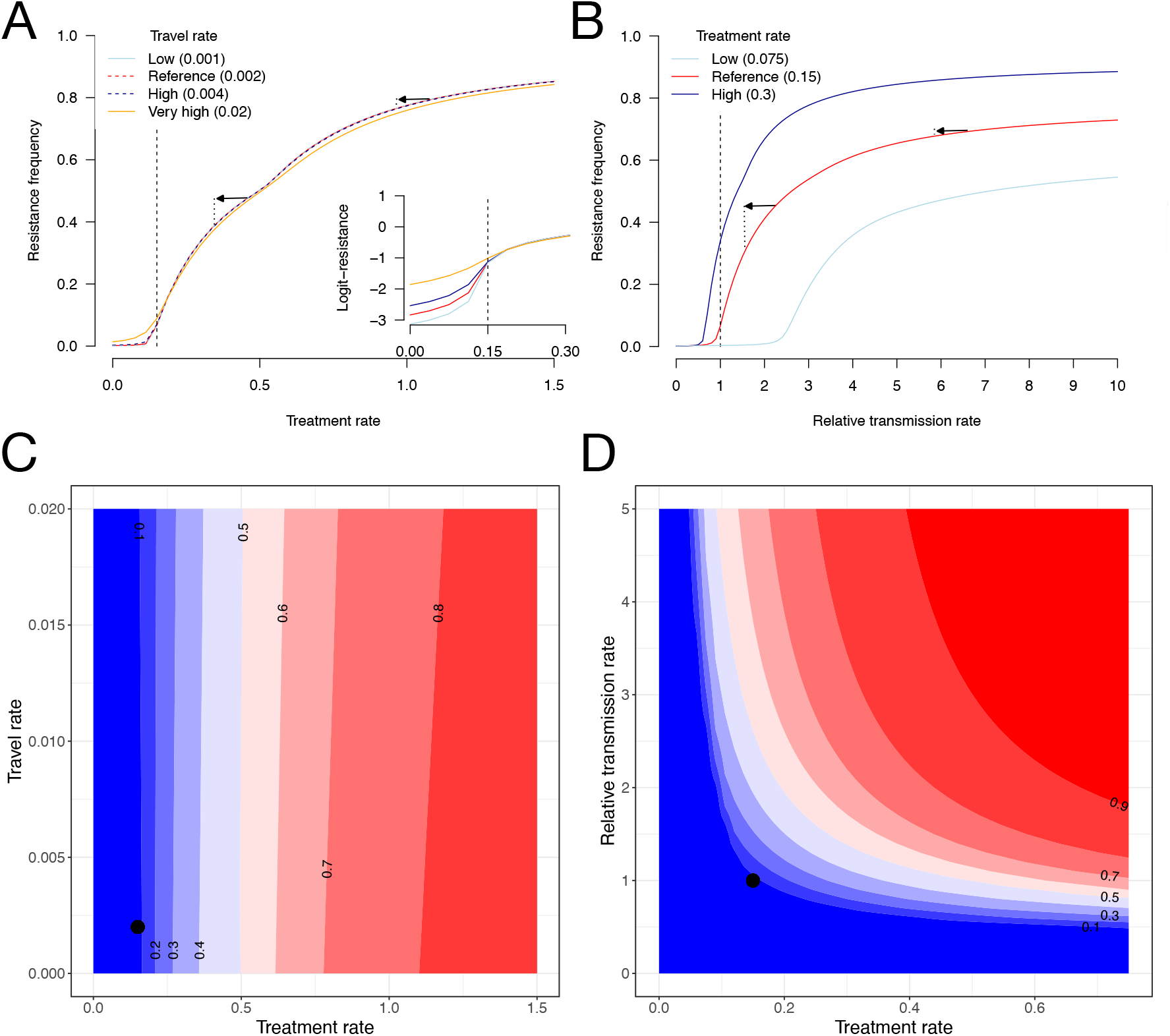
Sensitivity of the frequency of resistance to the rates of treatment, transmission and travel. **A**, the frequency of resistance as a function of the treatment rate, shown for four values of the travel rate. The dashed vertical line is the reference value. The inset shows the logit-transformed resistance frequency (log_10_(*p*_*R*_/(1 − *p*_*R*_)) specifically in the low-treatment region, highlighting the impact of travel in these conditions. **B**, the frequency of resistance as a function of the relative transmission rate, for three values of the treatment rate. On panels **A & B**, the arrows highlight the effect of a change in treatment rate or transmission on resistance frequency. **C, D** contour plots of the frequency of resistance as a function of treatment, transmission and travel rate. The points show the reference values. When unspecified, parameters are fixed to their reference value as given in Table 1.

#### Impact of the treatment rate

We predicted that a two-fold increase in the rate of antibiotic treatment would result in resistance increasing from the reference equilibrium value, 0.067 (predicted by the model), to 0.34 (a five-fold increase). A two-fold reduction in treatment would reduce resistance to 0.0031 (Fig. 4A, C, D).

#### Impact of the transmission rate

A two-fold increase in the transmission of *E. coli* increases resistance to 0.41 (a six-fold increase compared to 0.067), while a two-fold reduction reduces resistance to 0.0026 (a 26-fold decrease compared to 0.067) (Fig. 4B, D). Interestingly, the saturating shapes of the treatment-resistance and transmission-resistance relationships (Fig. 4A, B) imply that for the same baseline level of resistance, reducing transmission or treatment rate could have a small or large impact on the frequency of resistance (arrows on Fig. 4A, B).

#### Impact of the travel rate

In contrast, varying the rate of travel had very little impact on the equilibrium frequency of resistance (Fig. 4A, C). The only discernible impact was when the treatment rate is very small, in which case resistance is counter-selected and the equilibrium frequency of resistance is therefore proportional to travel (Fig. 4A inset). In population genetics terms, in this parameter region resistance is maintained at a selection-migration balance.

#### Negative frequency-dependent selection on resistance

We investigated the frequency-dependent selection (Harmand et al., 2019) on resistance generated by our model that includes a combination of mechanisms stabilizing resistance at an intermediate frequency. Our model generates negative frequency-dependent selection (NFDS), as shown by the slope of the selection coefficient on resistance as a function of its frequency in Figure 5, which results in the coexistence of resistant and sensitive strains. The equilibrium frequency of resistance is where the curve crosses the x-axis. The selection coefficient on resistance first rapidly declines, before decreasing as the frequency of resistance increases. The initial phase of strong positive “selection” is actually driven by travel and importation of resistance from abroad. This initial increase is faster when the travel rate is high. The second phase of decline depends on the treatment and transmission rates. Increasing the treatment or the transmission rate strengthen selection in favor of resistance at large resistance frequency and consequently increases the equilibrium frequency of resistance (Fig. 5A,C).

**Figure 5:**
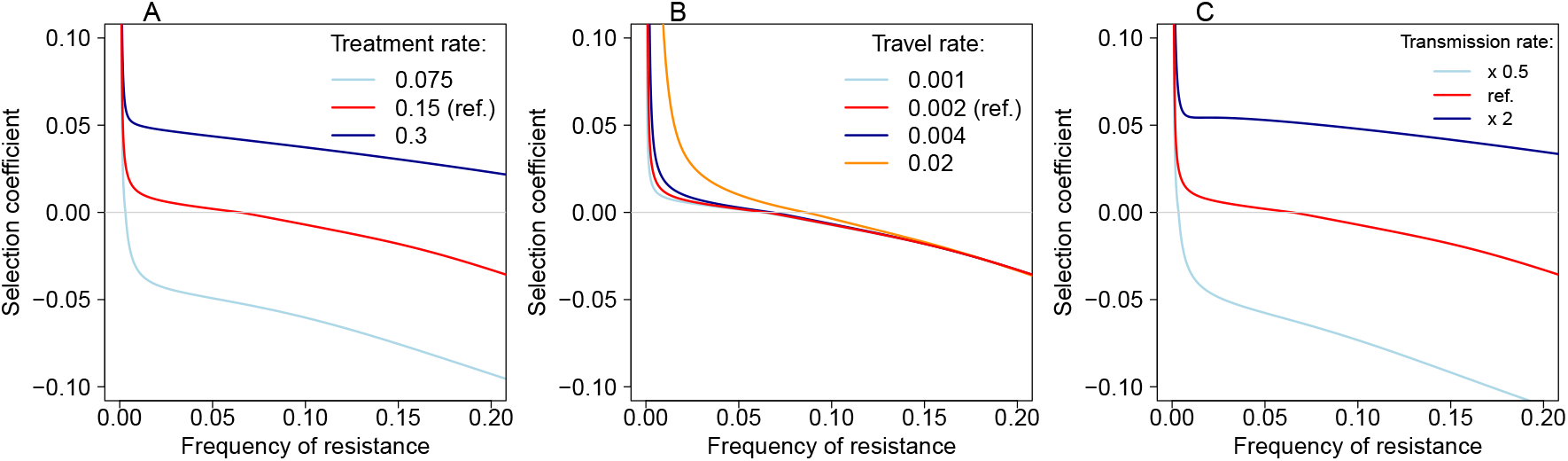
Effect of the treatment rate (A), travel rate (B) and transmission rate (C) on frequency-dependent selection on resistance. Frequency-dependent selection is shown by the selection coefficient on resistance as a function of resistance frequency. The selection coefficient is measured as *s*_*R*_ = *d/dt*(log(*p*_*R*_/(1 − *p*_*R*_)).

Modeling both a colonizer and a persistent strategy strengthens NFDS compared to a model assuming a single generalist strategy with the same average fitness (Fig. 6A). We further found that when both a colonizer and a persistent strategy are implemented, resistance is associated with the persistent strategy (Fig. 6B), while sensitivity is associated with the colonizer strategy (not shown). These associations increase when differentiation along the persistence-colonization trade off increases (Fig. 6B). Overall, the NFDS generated by the occurrence of differentiated strategies on the colonization-persistence trade-off, and the non-random association of resistance with these strategies, favor the coexistence of resistant and sensitive strains in the community.

**Figure 6:**
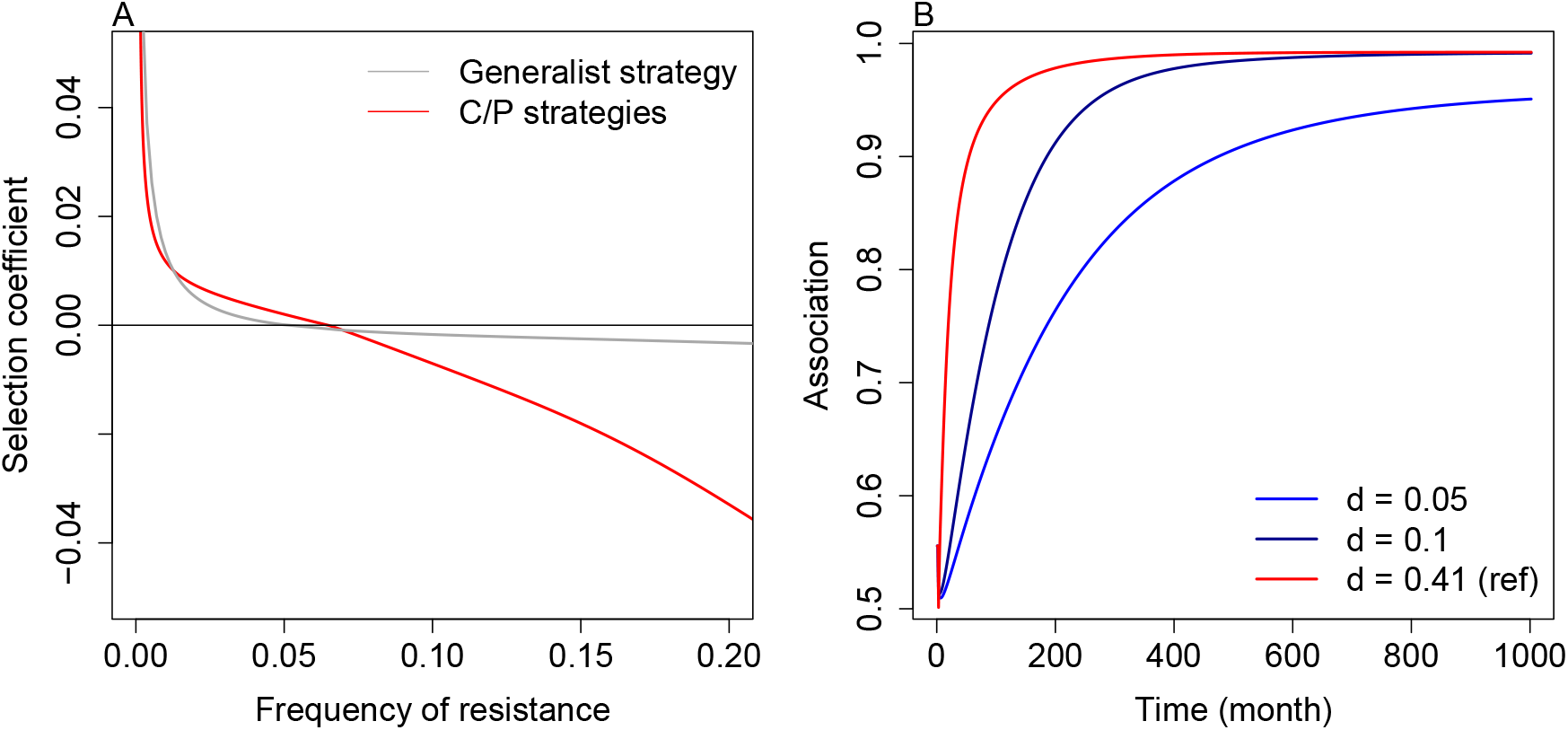
Effect of niche differentiation along the colonization-persistence trade-off on negative-frequency-dependent selection (NFDS). Frequency-dependent selection is shown by the slope of the selection coefficient on resistance as a function of resistance frequency. A: Comparison of NFDS produced by the (main) model with both a colonizer and a persistent strategy and that of a model with a single generalist strategy with the same fitness. In the generalist model, resistance costs (*P*_*R*_) were inferred similarly to the main model. B: Association of resistance with the persistent strategy in a model assuming differentiated strategies along the persistent/colonization trade-off. The association is measured as the proportion of *R*_*P*_−carrying hosts among all *R*-carrying hosts. Differentiation d is measured as departure from the mean strategy: 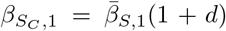 and 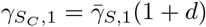 for the colonizer and opposite (− *d*) for the persistent. Other parameters as in Table 1.

## 4 Discussion

In Europe, the stabilization of the frequency of resistant *E. coli* at a low level after a rapid increase in the early 2000s remains largely unexplained (Emons et al., 2025). Empirical and theoretical studies have pointed to several mechanisms that favor resistance carriage and coexistence of resistant and sensitive strains in the community (Blanquart, 2019; Rahbé et al., 2024). However, how these mechanisms contribute to the observed dynamics remain largely unknown. Here,we proposed a data-informed epidemiological model that leverages i) a large dataset on ESBL-driven antibiotic resistance in healthy infants ii) a detailed microbiological characterization of samples and iii) our current knowledge of the ecology and epidemiology of *E. coli*, to gain a mechanistic understanding of the evolution of ESBL-driven antibiotic resistance in the community in France.

It has repeatedly been reported that returning from South and Southeast Asia is a risk factor for carriage of resistant strains of Enterobacterales. Although travel has been presented as an important contributor to resistance in high-income countries, and measures to limit the importation of resistance by travelers have been proposed (e.g., Reuland et al., 2016; Arcilla et al., 2017), the actual impact of travelers on local resistance had not been quantified. This impact not only depends on the rate of travel, but also on the dynamics of onward transmission of resistance by travelers and the eventual clearance of resistant strains. In our model including these phenomena, we show that importations have little effect on the local equilibrium ESBL frequency, except initially or when the local treatment rate is very low. At the current or higher treatment rates, a 10-fold increase in the rate of travel would barely change the equilibrium frequency of resistance in the focal community. Our results are in line with another recent mathematical model (Rahbe et al., 2025).

ESBL resistance must come with costs that counterbalance its advantage under treatment. Costs of antibiotic resistance have often been found in *in vitro* growth assays (e.g. Melnyk et al., 2015). However, experimental evidence on whether and how antibiotic resistance alters transmission is still missing (Andersson, 2006) and the costs in terms of epidemiological parameters have not previously been quantified. Here, we modeled and inferred four costs of resistance: two costs on transmission to empty and colonized hosts, and two costs on clearance in single- and co-colonized hosts. We could only precisely estimate the costs of transmission to colonized hosts and clearance in co-colonized hosts, which corresponds to the most frequent ecological context in which ESBL *E. coli* are found. These costs were found to be relatively low (−14% and +23% on transmission and clearance, respectively).

We also found that resistant strains transmit as much from single-colonized hosts as from mixed-carriage hosts (*m*_*x*_ close to 1), which is consistent with our data, where resistant strains reached similar density when they are alone in their host as when they are co-carried with sensitive strains. The model further implies that ESBL strains cannot persist in the community if resistance transmission from mixed-carriage hosts is lower than from hosts carrying the resistant strains only.

Overall, the relatively small cost of resistant strains in transmission in spite of substantially lower density could be explained if the relationship between within-host density and transmission rate is sublinear, as is the case for other pathogens where this relation was characterised (Fraser et al., 2007; Duong et al., 2015; Marc et al., 2021). Such relationship could be determined for ESBL resistance from household studies, but this has not yet been done to our knowledge (Arcilla et al., 2017; Perez et al., 2025).

In our model, the frequency of ESBL is strongly determined by the rate of treatment with beta-lactams. The relationship between the rate of treatment and the equilibrium level of resistance was quite progressive, in contrast to most previous theoretical work (Blanquart, 2019; Davies et al., 2019). Negative-frequency dependent selection (NDFS) is key to promote coexistence between resistant and sensitive strains (Davies et al., 2019). Our model with biologically plausible structure and parameters was able to generate NFDS and reproduce the progressive use-resistance relationship.

An important finding is that the variability in carriage duration, here modeled with a twice longer carriage duration for strains of phylogroup B2, was the most important factor favoring coexistence. Resistance preferentially evolves on the genotype with long duration of carriage in our model (Fig. S8) (Lehtinen et al., 2017). This prediction was also verified to some extent in our data: typing some of the resistant strains isolates from these data showed that ESBL resistance was mostly confined to ST131 (phylogroup B2) (Birgy et al., 2016), a sequence type with remarkably long duration of carriage (Johnson et al., 2022). Thus, the heterogeneity in duration of carriage must be modeled to properly reproduce qualitative patterns in data. Other factors do not generate much coexistence. The clearance-recolonization dynamics, which we modelled in line with available data (Cotto et al., 2023), is poorly conducive to coexistence (Fig. S6). Lastly, travel had a very limited role in stabilising coexistence: in spite of strong differentiation between France and Southeast Asia, the rate of travel was too small to significantly impact equilibrium levels of resistance.

Nevertheless, the observed coexistence and the NFDS were less strong in our model than in data. Among the factors that we did not include in the model, the structure in hospital-community and the acquisition of ESBL strains in hospitals may not play an important role: in broad terms, hospitalization has analogous consequences to travel. In France, there are an estimated 10M hospital stays per year. If about half of individuals return from hospital stays colonized by ESBL, the impact of hospitalization is equivalent to the impact of travel at a rate of 0.02 per month (10-fold greater rate), which is negligible. Moreover, in our data, infants with a recent hospital stay did not have particularly elevated levels of resistance (Table S1). Other unobserved sources of population structure could play a role in coexistence, but this would imply that a major risk factor for ESBL carriage has been overlooked in the many previous studies. The last and most plausible explanation in our view is that the distribution of carriage duration is variable at a finer phylogenetic unit (i.e. across ST). For lack of better data, we modelled a very simple distribution of carriage duration reflecting a two-fold longer carriage for B2 phylogroup vs. non-B2 phylogroup as in Swedish infants (Östblom et al., 2011). The true value of this ratio was larger (4-fold) in another study (Johnson et al., 2022). Modelling this distribution in more details may lead to a more progressive use-resistance relationship (Lehtinen et al., 2017), stronger NFDS, and should therefore be an important focus for future work.

We also found that increasing transmission increases the prevalence of resistance in the community. Importantly, the effect of transmission is nearly as strong as that of the rate of antibiotic treatment. This finding is consistent with a global analysis that found that factors associated with poor sanitation, but not antibiotic use, are associated with antibiotic resistance in *E. coli* (Collignon et al., 2018), and other recent modelling work (Rahbe et al., 2025). Transmission favors colonization by resistant strains during and immediately after treatment, when competition by sensitive strains is released (Blanquart et al., 2018; Davies et al., 2019). In untreated hosts, increasing transmission similarly favors resistant and sensitive strains without affecting their relative fitness (assuming a fixed cost of resistance). Without treatment, sensitive strains increase in frequency by out-competing resistant strains in the clearance-recolonization dynamic of untreated hosts. Explicitly modeling treatment structure is necessary to capture how transmission favors resistance (Blanquart et al., 2018) (a model without such structure predicts that transmission disfavors resistance− not shown).

Our dataset, in spite of its very large size, focus on healthy individuals, and detailed information, has some limits. We focused on healthy infants aged less than two years and use these data to model the community as a whole. We note that most studies focusing on resistance in the gut have to resort to convenience sampling, e.g. infants, pregnant women, hospital patients. Several results support that the epidemiology of *E. coli* resistance and sensitivity weakly depends on the age of the hosts. Children are very rapidly colonized by *E. coli* after birth (Palmer et al., 2007). The equilibrium ESBL frequency in our data was similar to that found in other age groups in the Paris area (Nicolas-Chanoine et al., 2013). Data from invasive infections collected by the European Center for Disease Prevention and Control (ECDC) also does not support a strong and consistent structure across age classes for *E. coli* ESBL resistance (Fig.S11). Lastly, the patterns in the data that inform our inferences are robust to examining infants, children or adults. History of treatment and travel as main risk factors for resistance are well established (e.g. Birgy et al., 2016), as is the fact that resistant most of the time co-occurs as a minority strains within hosts (Ruppé et al., 2013; de Lastours et al., 2016). Another potential limitation of our data is that children undergoing antibiotic treatment within 7 days prior to the sampling were excluded from the survey. These children potentially carry ESBL at higher frequency and density (Ruppé et al., 2013; de Lastours et al., 2016), which would lead to underestimating the frequency of ESBL in our focal community and the prevalence of individuals colonized only by resistant strains. This underestimation is likely to be weak, as given the observed rate of treatment we would miss only nine infants with this exclusion criterion. Accordingly, the ESBL frequency in our data is consistent with other studies (e.g. Gagliotti et al., 2011; Bezabih et al., 2021).

Secondly, while we carefully crafted our model to include all plausible processes affecting antibiotic resistance, some of the assumptions could be discussed. We did not specifically model individuals who both travelled and used antibiotics (N=7). It is unlikely that modeling these individuals would have impacted our results, given the limited role of travel that we found and the model constrains some travelers to come back with ESBL. We did not include hospitalization as a risk factor in our model, but as discussed above hospitalization was not a risk factor in our data, and in theory cannot play a very important role. We assumed that co-colonized hosts carry and transmit twice as much bacteria than single-colonized ones. Models sometimes alternatively assume that the transmission rate is the same for single- and co-colonized hosts (Alizon et al., 2013). The predictions of our model were not affected by this assumption (Fig.S3). Lastly, our model allows only weak competitive release of the resistant strain: treatment leads to clearance of the sensitive strain, and the resistant strain does not transmit more when it is alone in the host. Empirical data suggest that treatment clears a large fraction of the whole microbiota. This could allow a stronger increase in the density of resistant strains than modelled here (Ruppé et al., 2013; Niehus et al., 2020). Our predictions remain unchanged when assuming a 5-fold increase in transmission of resistance upon treatment (Fig.S4).

Our findings have several implications. The most important drivers of local resistance frequency were antibiotic use in the community and transmission, not the importation of resistant strains from Southeast Asia. Coexistence was mainly stabilized by the interaction with another locus determining the duration of carriage (Lehtinen et al., 2017). This association has several important consequences. First, it is difficult to measure a cost of resistance on strain clearance because resistance is associated with strains with long duration of carriage. To directly measure this cost, it is necessary to control for the impact of the genetic background on carriage duration (Krishna et al., 2025). Second, an intervention to decolonize resistant strains of *E. coli* should be very efficient: indeed, if resistant strains are associated with “slow-turnover” strains with long duration of carriage and low colonization rates (Morel-Journel et al., 2023), then removing these strains would severely reduce their fitness. Thirdly, novel resistances might primarily emerge first on travellers, second on long-duration of carriage genomic backgrounds. Strains with long duration of carriage could be particularly targeted for surveillance. It would thus be necessary to quantify the distribution of residence times of bacterial strains in commensalism, and understand what factors maintain this diversity, to fully understand resistance evolution.

To conclude, using one of the largest cohorts studying ESBL resistance in the community in healthy individuals, together with detailed and informed modeling of competition between sensitive and resistant strains–a critical point that has been overlooked by some previous studies–we understood mechanistically the evolution of resistance in the community in France. This points to around 10-20% costs of resistance on transmission and clearance, a major role of selection by local antibiotic use and transmission, and variability in the duration of carriage as the main factor stabilizing sensitive and resistant strains. This opens perspectives for the quantitative study of resistance evolution, most interestingly testing our prediction on the sublinear relationship between strain density and transmission (for example in household studies); and improved characterization of the duration of carriage of commensal bacterial strains.

## Data Availability

All data analyzed in the present work will be available at 10.5281/zenodo.18480481. The data are uploaded but not yet public. We will publish them after completing the revisions of the paper.

## 5 Acknowledgments

This work was funded by the CNRS Momentum grant to FB and the ERC StG 949208 to FB.

We are grateful to the INRAE MIGALE bioinformatics facility (MIGALE, INRAE, 2020. Migale bioinformatics Facility, doi: 10.15454/1.5572390655343293E12) for providing computing resources.

Data for Figures 2 and S11 from The European Surveillance System—TESSy, provided by Austria, Belgium, Czech Republic, Germany, Denmark, Spain, Finland, France, Hungary, Ireland, Italy, Netherlands, Norway, Poland, Portugal, Sweden, Slovenia, United Kingdom, and released by ECDC. The views and opinions of the authors expressed herein do not necessarily state or reflect those of ECDC. The accuracy of the authors’ statistical analysis and the findings they report are not the responsibility of ECDC. ECDC is not responsible for conclusions or opinions drawn from the data provided. ECDC is not responsible for the correctness of the data and for data management, data merging and data collation after provision of the data. ECDC shall not be held liable for improper or incorrect use of the data.

## 6 Supplementary material

### S1 Statistical analysis

#### Summary of the full data set

3443 rectal samples from French children aged 6-24 months have been collected from 2010 to 2018, corresponding to 3417 different individuals. Sampling was associated with a questionnaire. Information so gathered is fully described in (Birgy et al., 2016).

Birgy et al. (2016) further analyzed the risk factors associated with ESBL carriage (based on samples obtained during the 2010-2015 period). In particular, a multivariate analysis identified home care, the use antibiotics and the travel history within three months prior to sampling as the main risk factors related to ESBL carriage. Furthermore, univariate analyses identified travel to Oceania/Asia as the only region associated with a significant increase in the risk of ESBL carriage (Birgy et al., 2016).

In the present study, we focused on the risk factors related to ESBL carriage corresponding to i) travel to Asia/Oceania, hereafter Southeast Asia [SEA], and ii) the use of antibiotics, both within three months prior to sampling (Table S1). We extended the analyses of Birgy et al. (2016) to the period 2010-2018 and found similar results (Table S2).

**Table S1:** Contingency table for the number of occurrences of each risk factor considered in the model.

|  |  | Used antibiotics three month before sampling |  |
| --- | --- | --- | --- |
|  |  | 0 | 1 |
| Traveled to SEA three | 0 | 2150 | 1268 |
| month before sampling | 1 | 12 | 7 |

**Table S2:**
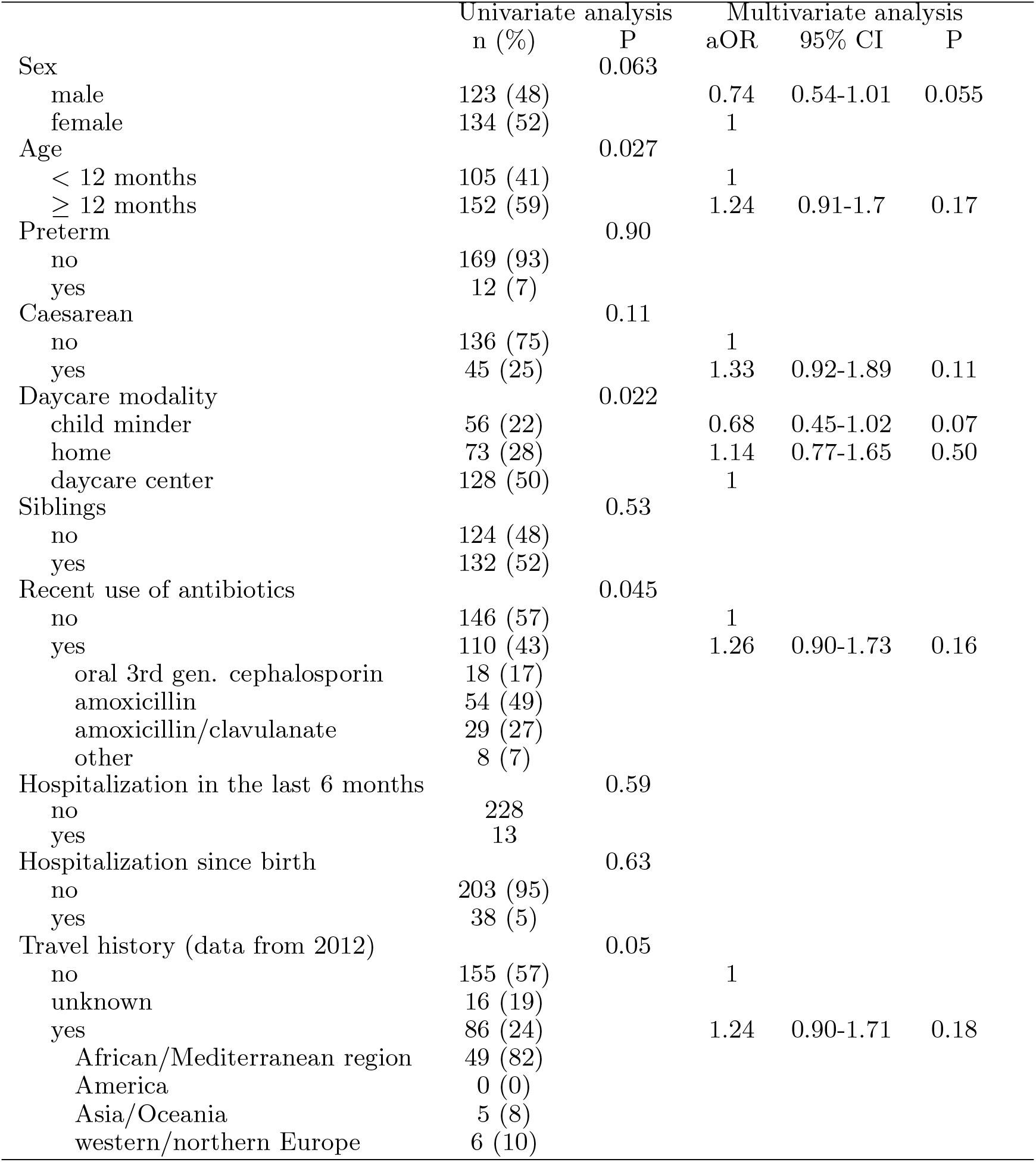
Analysis of risk factors associated with ESBL carriage (extended from Birgy et al. 2016 to the period 2010-2018); overall samples N = 3443, carriage of ESBL = 257 (7.4%). The univariate analysis was performed using *χ*-square tests. For the multivariate analysis, we kept only the most significant factors of the univariate analyses (P*<*0.2). Children who did not travel were pooled with those who travelled to western/northern Europe. In the multivariate analysis, we considered only the children with information on travel history (2012-2018).

### S2 Density of resistant and sensitive E. coli strains in mixed-carriage hosts

Among the above samples (years 2014-2016), 301 were further analyzed to evaluate the total density of Enterobacterales (EB) and the density of ESBL-carrying *E. coli* strains. Density has been estimated using dilution counts (by a factor 10^−1^ to 10^−6^) on selective media (Drigalski for Enterobacterales and ESBL-selective). There was five samples where no EB were detected.

We further assumed that the number of bacteria in each sample followed a Poisson distribution. The Maximum Likelihood estimator for the number of bacteria is then

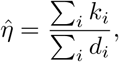

where *d*_*i*_ is the dilution factor and *k*_*i*_ is the number of bacteria counted at dilution *d*_*i*_.

For each sample where EB were found, we then compared two models using a AIC criterion (i.e. ΔAIC ≥ 2): i) *η*_*EB*_ = *η*_*ESBL*_ (all EB carry ESBL genes), ii) *η*_*EB*_ ≠ *η*_*ESBL*_ (some EB carry ESBL genes). The results are presented on table S3.

**Table S3:**
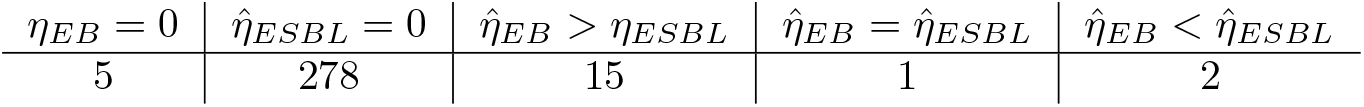

| $\eta_{EB} = 0$ | $\hat{\eta}_{ESBL} = 0$ | $\hat{\eta}_{EB} > \eta_{ESBL}$ | $\hat{\eta}_{EB} = \hat{\eta}_{ESBL}$ | $\hat{\eta}_{EB} < \hat{\eta}_{ESBL}$ |
| --- | --- | --- | --- | --- |
| 5 | 278 | 15 | 1 | 2 |

We found five samples without EB cells (Tab. S3). In most samples with ESBL strains, ESBL strains co-occurred with sensitive strains. We found three samples where all EB carried ESBL, including two samples in which the number of EB cells carrying ESBL was estimated to be larger than the total number of EB cells, probably due to measurement error. Figure S1 shows the distribution of the relative ESBL density. We found that in most cases, ESBL density represents more than 1% of the total EB density (30% quantile at 1% and mean at 17%). In our likelihood calculation, we considered that hosts with 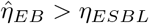 belonged to type “RS”, and we used the number of uncolonized hosts *η*_*EB*_ = 0 (see main text).

**Figure S1:**
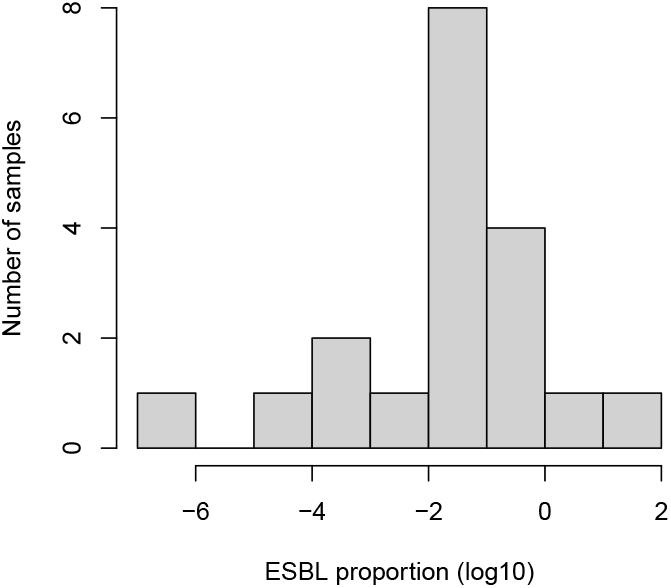
Distribution of the relative ESBL density

### S3 Parameters

We provide here the methods used to obtain the parameter values that are not inferred with the model.

#### Treatment rate

In the focal data set, on average 37% (min 33%, max 43% depending of the year) of children received antibiotics within three months before sampling. The number of children starting an antibiotic treatment during a given time period follows a Poisson process. The elapsed time *T* between the end of the treatment and the sampling follows an exponential law of parameter *τ*. The probability that the sampling occurs during a three-month period after treatment is *P* (*T* ≤ 3) = 1 − *e*^−3*τ*^ = 0.37. Solving for this equation, we find *τ* = 0.15 *m*^−1^. The rate at which the event “sampling occurs within three months after treatment” occurs is equal to the rate of occurrence of “starting an antibiotic treatment”, such that *τ* is the treatment rate.

#### Duration of the treatment

The typical duration of antibiotic treatments is one week (Spellberg and Rice, 2019), corresponding to a monthly rate of 4 *m*^−1^.

#### Antibiotic efficiency

We used results from Paterson et al. (2016) who investigated the optimal antibiotic concentration to cure bacterial infection. Using their typical values for antibiotic concentration (23 *µg*.*L*^−1^) and sensitive strains in their equation 5, we find an approximate death rate due to antibiotics of 8.6 *d*^−1^ in their original scaling, corresponding to 266 *m*^−1^.

#### Post-treatment period

By choosing *ω*_3_ = 1/3 *m*^−1^ the mean time spent in the compartment T3 is three months.

#### Travel rate

In the focal data set, the proportion of children traveling to Southeast Asia varied between 0 and 2% per year, with mean 0.7% per year. The travel rate is calculated as the treatment rate. It is the solution of *P* (*V* ≤ 3) = 1 − *e*^−3*ν*^ = 0.007, such that *ν* = 0.002 *m*^−1^.

#### Duration of travel

Most studies estimated the mean travel time to SEA is about 20 days (e.g. Ruppé et al., 2015; Arcilla et al., 2017), corresponding to a rate of return from travel of 1.5 *m*^−1^.

#### Post-travel period

As for the compartment T3, we chose *ψ*_3_ = 1/3 *m*^−1^ such that the mean time spent in this compartment is three months.

#### Force of infection during travel

Previous studies showed that 30 to 70% of travelers to SEA are colonized by ESBL strains during travel (Paltansing et al., 2013; Ruppé et al., 2015; Kantele et al., 2015; Reuland et al., 2016; Arcilla et al., 2017; Woerther et al., 2017). We assumed that about 50% of the travelers are colonized during a 20-day travel (Ruppé et al., 2015). We further assumed that the force of colonization of resistant strains did not depend on their colonization type: 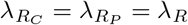.

A simple model of infection can inform on the force of colonization while traveling. Consider a cohort of individuals that travel to Asia during 20 days. None of them initially carry MRE. The dynamics of MRE carriage during travel is given by: *dX*_*R*_[*t*]*/dt* = *λ*_*R*_(1 − *X*_*R*_[*t*]) (neglecting clearance during travel) where *X*_*R*_ is the density of travelers carrying MRE during travel, *λ*_*R*_ is the force of colonization during travel (assumed to be independent of *X*_*R*_ and *t*). Solving for the differential equation with *X*_*R*_[0] = 0, we obtain 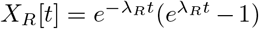. Further using *t* = 20 and looking for *X*_*R*_[20] = 0.5*N* and solving for *λ*_*R*_, we obtain *λ*_*R*_ ≈ 0.035 ≈ *d*^−1^ 1.1 *m*^−1^

No information is available concerning the force of colonization by sensitive strains during travel. As a baseline, we assumed that *λ*_*S*_ ≈ *λ*_*R*_.

#### Clearance and colonization rates of sensitive strains

We estimated these parameters using an external dataset on the turnover of *E. coli* strains in 127 Swedish infants followed longitudinally from birth (Nowrouzian et al., 2003, 2005; Östblom et al., 2011). The infants were followed at 3 days, 1 week, 2 weeks, 4 weeks, 2 months, 6 months, 12 months after birth. At each visit, *E. coli* colonies were isolated on Drigalski agar from faecal samples. As detailed in Östblom et al. (2011), one to six colonies with different morphologies were isolated, typed with Random Amplified Polymorphic DNA (RAPD) profiling and genotyped with several PCRs. Here, we used the phylogroup of these strains obtained from Clermont typing (Clermont et al., 2000). Phylogroup B2 was associated with a longer duration of carriage than others (Nowrouzian et al., 2005; Östblom et al., 2011). Thus, we identified “colonizer” and “persistent” types to non-B2 and B2 strains. We used a Bayesian approach to estimate the colonization and clearance rates depending on both the colonization status of the host and on the phylogroup of the strains.

##### Overview of the model fitted to longitudinal data

We described the strain turnover within infants by a continuous time, discrete-state Markov chain model. We defined the strains as 0 (0 ≡ C ≡ non-B2) or 1 (1 ≡ P ≡ non-B2). We considered as variables the frequency of empty hosts, hosts colonized by a single strain, and hosts colonized by two strains. Colonization by more than two strains only occurred twice, and these two timepoints were removed from our analysis. There were therefore six variables representing the system: 1 for empty hosts, 2 for the single colonizations, 3 for the co- colonizations. Transitions between colonization states occurred in three ways:

- colonization by one of the strains happening at rates *λ*_0_, *λ*_1_, *λ*_00_, *λ*_01_, *λ*_10_, *λ*_11_. These rates account for both the transmission rate and the frequency of the colonizing strain in the community. For co-colonization, the first index represents the strain already present in the host, and the second index represents the colonizing strain.
- clearance of one of the strains happening at rates *µ*_0_, *µ*_1_, *µ*_00_, *µ*_01_, *µ*_10_, *µ*_11_. For clearance of co-colonized individuals, the first index represents the strain that remains, and the second index represents the strain that is cleared.
- direct replacement of one of the strains by another happening at rates *ν*_01_, *ν*_10_. The second index represents the strain that replaces.

A 6 × 6 transition matrix **A** described the rates of transitions between host colonization states. The probability of a specific observed change from state *i* to state *j* in an infant, over an interval of duration Δ*t*, was given by exp (**A**Δ*t*)_*i,j*_. The probability of observed succession of states in an infant was the product of these probabilities. Finally, the likelihood–the probability of the dataset given the parameters–was given by the product of all infant probabilities.

Our aim was to use MCMC to estimate the posterior distribution of the 14 parameters:

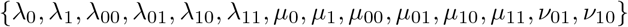

After initial fits of the model where strains were considered as all equivalent, we used exponential priors with means (in months^−1^):

- 0.18 *m*^−1^ for simple colonizations *λ*_0_, *λ*_1_
- 0.066 *m*^−1^ for co-colonizations *λ*_00_, *λ*_01_, *λ*_10_, *λ*_11_
- 0.078 *m*^−1^ for clearance in single colonization *µ*_0_, *µ*_1_
- 0.234 *m*^−1^ for clearance in co-colonizations *µ*_00_, *µ*_01_, *µ*_10_, *µ*_11_
- 0.00192 *m*^−1^ for replacements *ν*_01_, *ν*_10_

We ran an MCMC algorithm for 2 × 10^6^ iterations and retained only the last 1 × 10^6^ iterations to estimate the posterior distribution of parameters, after visually checking convergence of the algorithm.

##### Estimation of clearance rates

We used the mean posterior distribution of the six clearance rates (see below for the transmission rates), which were estimated to:

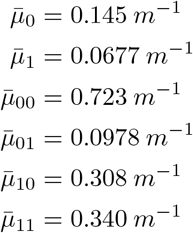

These parameters are related to the parameters of our model following:

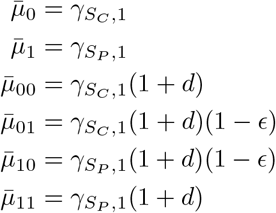

where *d* measures the additional clearance from co-colonized hosts (such that *γ*_*k*,2_ = *γ*_*k*,1_(1 − *d*)) (*k* ∈ {*S, P*}) and *ϵ* measures niche differentiation (see main text). We used a linear model to estimate the 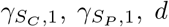 and *ϵ* (the four model parameters) from the mean of the posterior distributions using

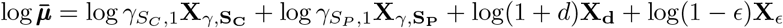

where 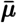 is the vector of the mean of the posterior for each of the clearance parameters, the **X**_*i,k*_ are binary vectors with value depending on whether 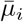 contains the corresponding factor *k*. The parameters 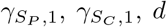 and *ϵ* can be directly obtained from the estimated coefficients of the linear model. Solving this linear model, we thus obtained 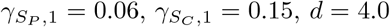 and *ϵ* = 0.65. These estimates result in the clearance parameters presented in Table 1.

##### Transmission rates to uncolonized hosts

We used the mean posterior distribution of the six transmission rates, divided by the frequency of strain 0 (colonizers) or 1 (persisters) in the community, to estimate the transmission rates:

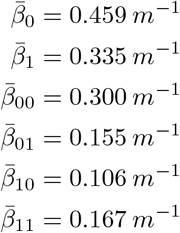

These parameters are related to the parameters of our model following:

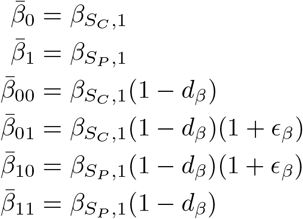

where *d*_*β*_ measures the reduced transmission to already colonized hosts and *ϵ*_*β*_ measures niche differentiation acting on transmission. We used a linear model to estimate the 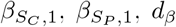 and *ϵ*_*β*_ (the four model parameters) from the mean of the posterior distributions using

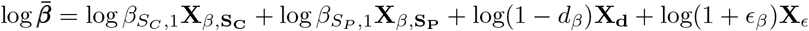

where 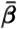 is the vector of the mean of the posterior for each of the transmission parameters, the **X**_*i,k*_ are binary vectors with value depending on whether 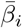 contains the corresponding factor *k*. The parameters 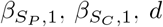 and *ϵ*_*β*_ can be directly obtained from the estimated coefficients of the linear model. Solving this linear model, we thus obtained 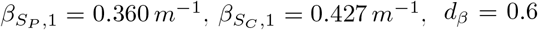. As the term corresponding to niche differentiation on transmission was not significant in the linear model, and in accordance with another study, we considered there was no niche differentiation on transmission (*ϵ*_*β*_ = 0.).

The inferred transmission rates in the Swedish cohort were not large enough to reach the observed prevalence of *E. coli* in our French cohort, perhaps because of differences in epidemiology between Sweden and France. Furthermore, these transmission rates together with the inferred clearance rates led to large difference in competitive ability between persisters and colonizers. We therefore modified the transmission rates to better fit our data. The transmission rates to uncolonized hosts were instead computed to ensure that the P- and C-types (roughly) have the same *R*_0_ and that the expected frequency of uncolonized hosts was (roughly) *S*^∗^ = 1.7 % as observed in our data. Precisely, we solved:

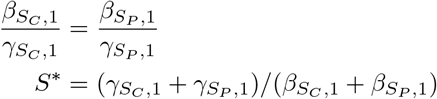

The second equation corresponds to a rough approximation for the expected frequency of un-colonized hosts. Solving these two equations for 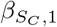, and 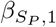, results in 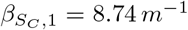 and 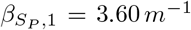 as presented in Table 1. We finally used the inferred effect of colonization on transmission rates value of, estimated at *d*_*β*_ = 0.6 to compute the values of 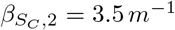 and 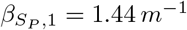. The final transmission parameters are presented in Table 1.

Finally, note that the data from the 127 Swedish infants did not support the mechanism of direct strain replacement: we estimated the mean posterior of these parameters to *ν*_01_ = 0.0055 *m*^−1^ and *ν*_10_ = 0.0055 *m*^−1^.

### S4 Metropolis-Hastings (MH) algorithm with adaptive sampling

In the following, we describe the MH algorithm used to obtain the MCMC chains.

Let be:

- *P*_*R,i*_ the vector of parameter values at iteration *i*
- *L*_*i*_ the likelihood of *P*_*R,i*_
- Σ_*P*_ the covariance matrix between parameters
- *U* the number of iterations to be performed between two updates of the covariance matrix. We used *U* = 500.

Initialization

- *P*_*R*,0_ a set of parameters drawn randomly in the respective prior distributions
- Σ_*P*_ a diagonal matrix with diagonal elements corresponding to the initial variances of the parameters (we used 10^−5^)

The algorithm then proceeds as follow

- if *i* is a multiple of *U*, we update the covariance matrix to optimize the exploration of the parameter space:
  – Determine the acceptance rate *a* since the last update as 1−*N* (Duplicate in {*P*_*R,k*_}_*k*∈[*i*−*U*+1,*i*]_)*/U*
  – Get the number *N*_*u*_ of unique elements in the chain {*P*_*R,k*_}_*k*∈[*i*−*U*+1,*i*]_.
  – if *N*_*u*_ = 1 (no proposal was accepted during the last *U* iteration), we reduce the covariance matrix to Σ_*P*_ = Σ_*P*_ */γ*, where we chose *γ* = 10.
  – if *a >* 0.5 (the acceptance rate is too large) we expand the covariance matrix to Σ_*P*_ = *γ* x Σ_*P*_.
  – if *a* ≥ 0.05 (the acceptance rate is too small), we calculate the covariance matrix Σ_*P,U*_ of the chain of unique elements in {*P*_*R,k*_}_*k*∈[*i*−*U*+1,*i*]_ and set Σ_*P*_ = Σ_*P,U*_ /(10 x *γ*)
  – if *a <* 0.05, we reduce the covariance matrix Σ_*P*_ = Σ_*P*_ */γ*
- else
  – proposal = *P*_*R,i*_ + *δ* with *δ* drawn in a centered multivariate normal distribution with covariance matrix Σ_*P*_
  – If each element of the proposal is within the non-zero interval of the prior distributions calculate the acceptance ratio as *α* = ℒ (*proposal*)/ℒ (*P*_*R,i*_) where ℒ denotes likelihood, draw a random value *x* in a Uniform distribution on [0, 1]. If *x < α* the proposal is accepted: *P*_*R,i*+1_ =proposal and *L*_*i*+1_ = ℒ (*proposal*), else the proposal is rejected: *P*_*R,i*+1_ = *P*_*R,i*_ and *L*_*i*+1_ = *L*_*i*_.
  – If an element of the proposal falls outside of prior, the proposal is rejected *P*_*R,i*+1_ = *P*_*R,i*_ and *L*_*i*+1_ = *L*_*i*_.

### S5 MCMC convergence analysis

We ran five independent MCMC chains with initial conditions sampled randomly in the prior distributions. The MCMC chains were ran for 250,000 iterations. The convergence of the MCMC chains was verified using functions (see below) provided in the coda R-package (Plummer et al., 2006).

**Table S4:**
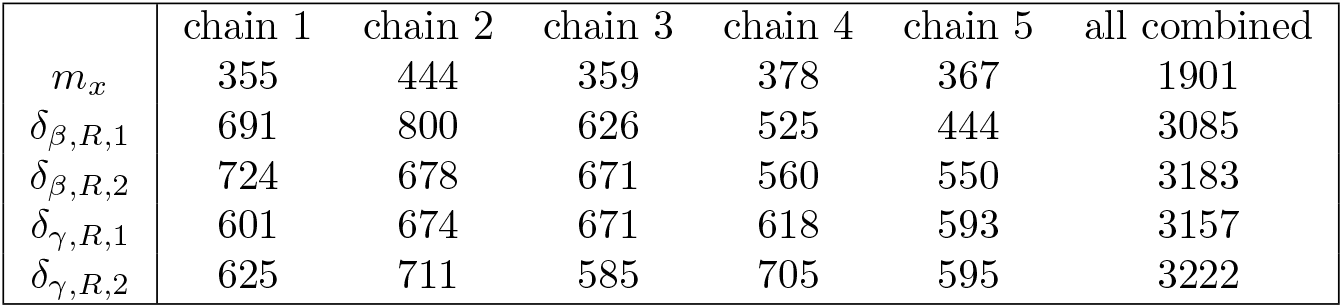
Sample size adjusted for the autocorrelation (rounded to the nearest integer)

|  | chain 1 | chain 2 | chain 3 | chain 4 | chain 5 | all combined |
| --- | --- | --- | --- | --- | --- | --- |
| $m_x$ | 355 | 444 | 359 | 378 | 367 | 1901 |
| $\delta_{\beta,R,1}$ | 691 | 800 | 626 | 525 | 444 | 3085 |
| $\delta_{\beta,R,2}$ | 724 | 678 | 671 | 560 | 550 | 3183 |
| $\delta_{\gamma,R,1}$ | 601 | 674 | 671 | 618 | 593 | 3157 |
| $\delta_{\gamma,R,2}$ | 625 | 711 | 585 | 705 | 595 | 3222 |

**Table S5:** Gelman and Rubin’s convergence diagnostic (implemented in R-package coda)

| | $m_x$ | $\delta_{\beta,R,1}$ | $\delta_{\beta,R,2}$ | $\delta_{\gamma,R,1}$ | $\delta_{\gamma,R,2}$ |
| --- | --- | --- | --- | --- | --- |
| point estimate (upper 95%CI) | 1.10 (1.01) | 1.01 (1.03) | 1.02 (1.04) | 1.01 (1.03) | 1.02 (1.05) |

**Figure S2:**
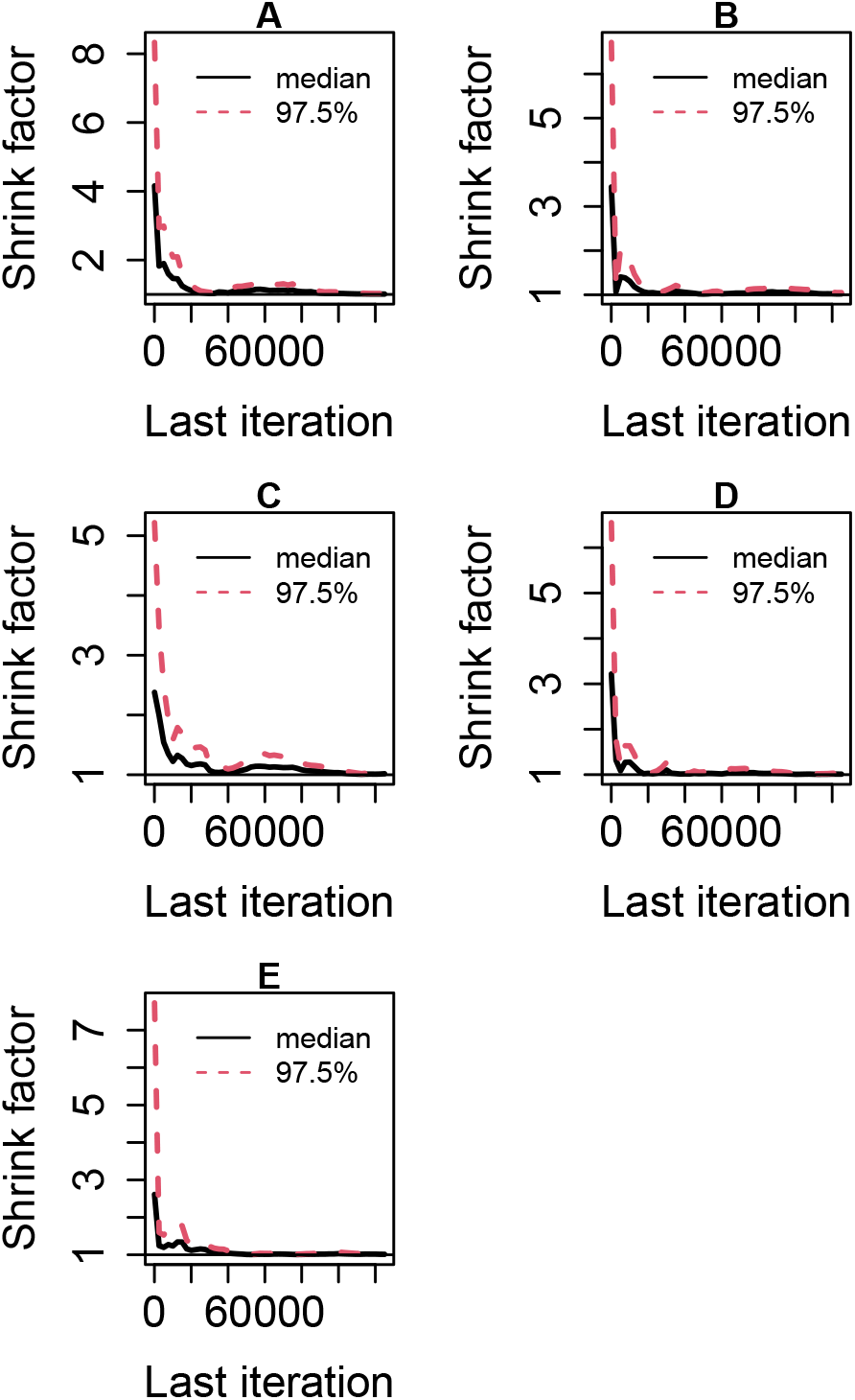
Gelman-Rubin convergence plots. A: *m*_*x*_, B: *δ*_*β,R*,1_, C: *δ*_*β,R*,2_, D: *δ*_*γ,R*,1_, E: *δ*_*γ,R*,2_

### S6 Sensitivity of the inference to the model assumptions

We investigated the sensitivity of our inference to two alternative scenarios. For brevity, we present only the posterior distributions obtained for each alternative scenario. Mostly, we found that our main results qualitatively hold. The main effects of the alternative scenarios considered here are on the estimate of the cost of resistance on transmission to uncolonized hosts, as we present below.

First, we assumed that the transmission rate of R-strains increases when hosts are under antibiotic treatment (Fig.S3). This scenario models strong competitive release during antibiotic treatment (e.g. Smith et al., 2021). Precisely, we assumed that, in the compartment *T* (hosts under treatment), 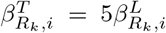, where *L* ∈ {*U, T*_3_, *SEA, SEA*_3_} and where *i* ∈ {1, 2} and *k* ∈ {*P, C*}. In this scenario, we obtained a relatively sharp estimate of a strong cost of resistance on transmission to uncolonized hosts (Fig.S3A), in contrast to the reference scenario (Fig. 3). Thus, assuming strong competitive release was counterbalanced by inferring a strong cost of resistance on transmission to uncolonized hosts.

Second, we modelled the case where transmission and clearance did not depend on density. Precisely, transmission and clearance rates from co-colonized hosts were the same as those from single-colonized hosts: *β*_*xx*_ = *β*_*x*_ and *γ*_*xx*_ = *γ*_*x*_, with *x* ∈ {*R*_*k*_, *S*_*l*_} and *k, l* ∈ {*C, P*}. The posterior distributions were similar to those obtained in the reference model (Fig.S4).

**Figure S3:**
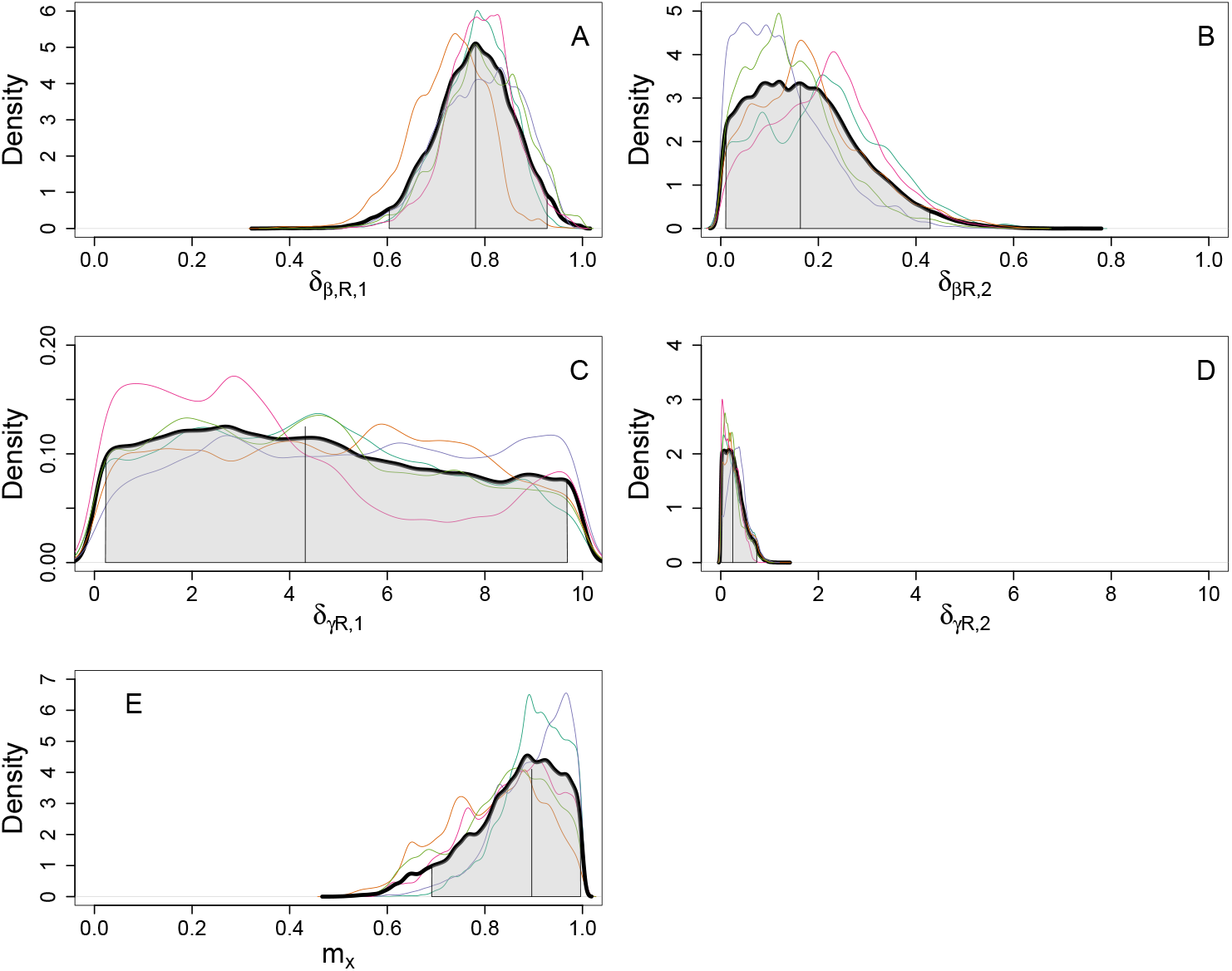
Posterior distribution under the assumption of strong competitive release during treatment. For all panels: colored curves represent the posterior distribution for each MCMC chain. The thick black curves is the posterior for all chains pooled. The vertical lines are the mean of the overall posterior distribution. Shaded area: 95% CI.

**Figure S4:**
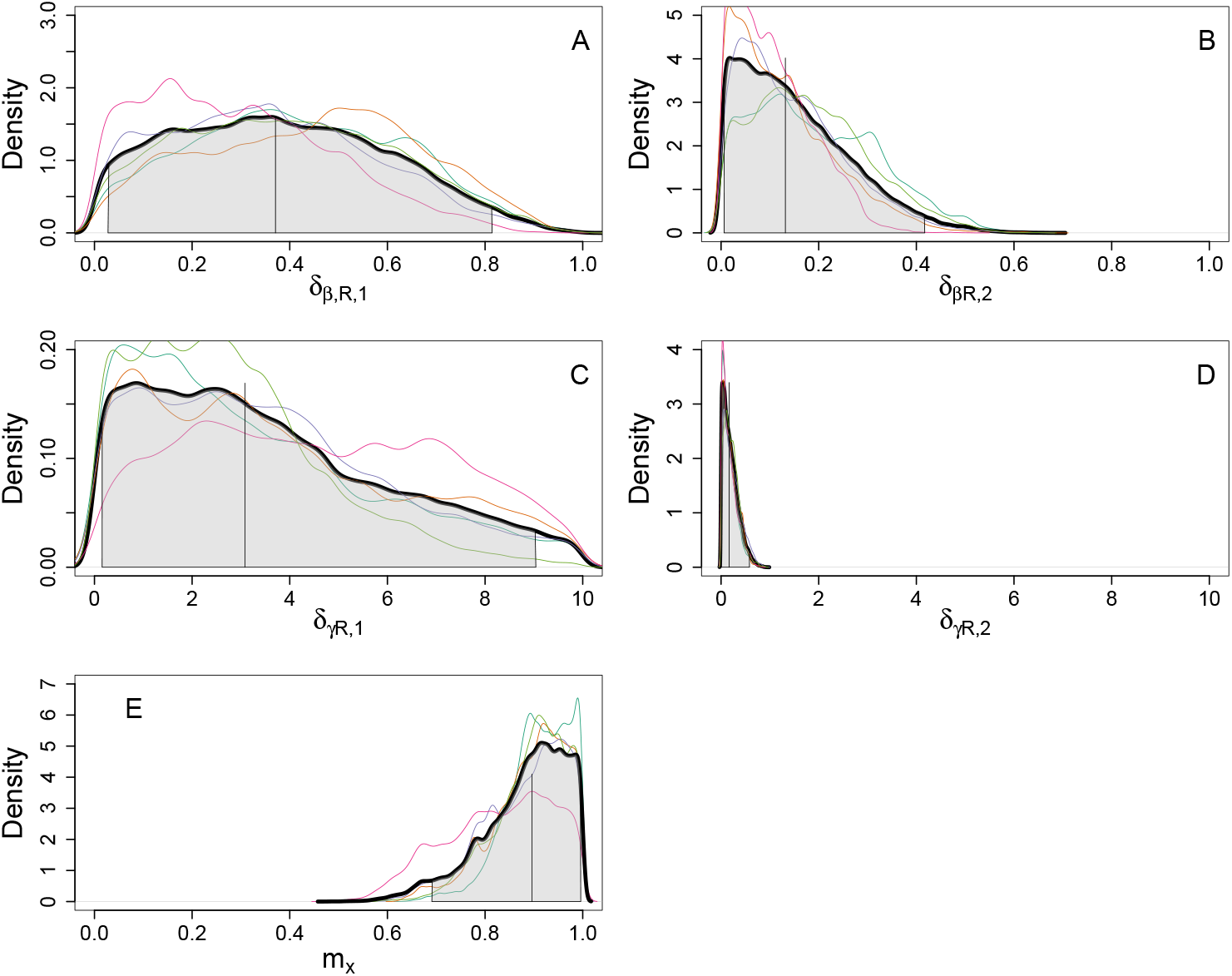
Posterior distribution under the assumption of density independent transmission and clearance rates. For all panels: colored curves represent the posterior distribution for each MCMC chain. The thick black curves is the posterior for all chains pooled. The vertical lines are the mean of the overall posterior distribution. Shaded area: 95% CI.

### S7 Model dynamics and comparison of the two-strain model with the four-strain model

#### Two-strain model

We initially considered the dynamics of resistant (R) and sensitive (S) strain, without genetic structuring. This model can be retrieved from the model presented in the main text (considering additional genetic structuring with persistent and colonizer strains) by assuming 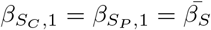 and 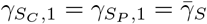. The results of the inference were similar to those we obtained with the main model (Fig. S5). However, the model allowed coexistence in a narrower range of parameters, especially for treatment rate (Fig. S6). Moreover, in the two-strain model the dynamics to equilibrium was very slow, contrasting with the observed dynamics (Fig. S7).

**Figure S5:**
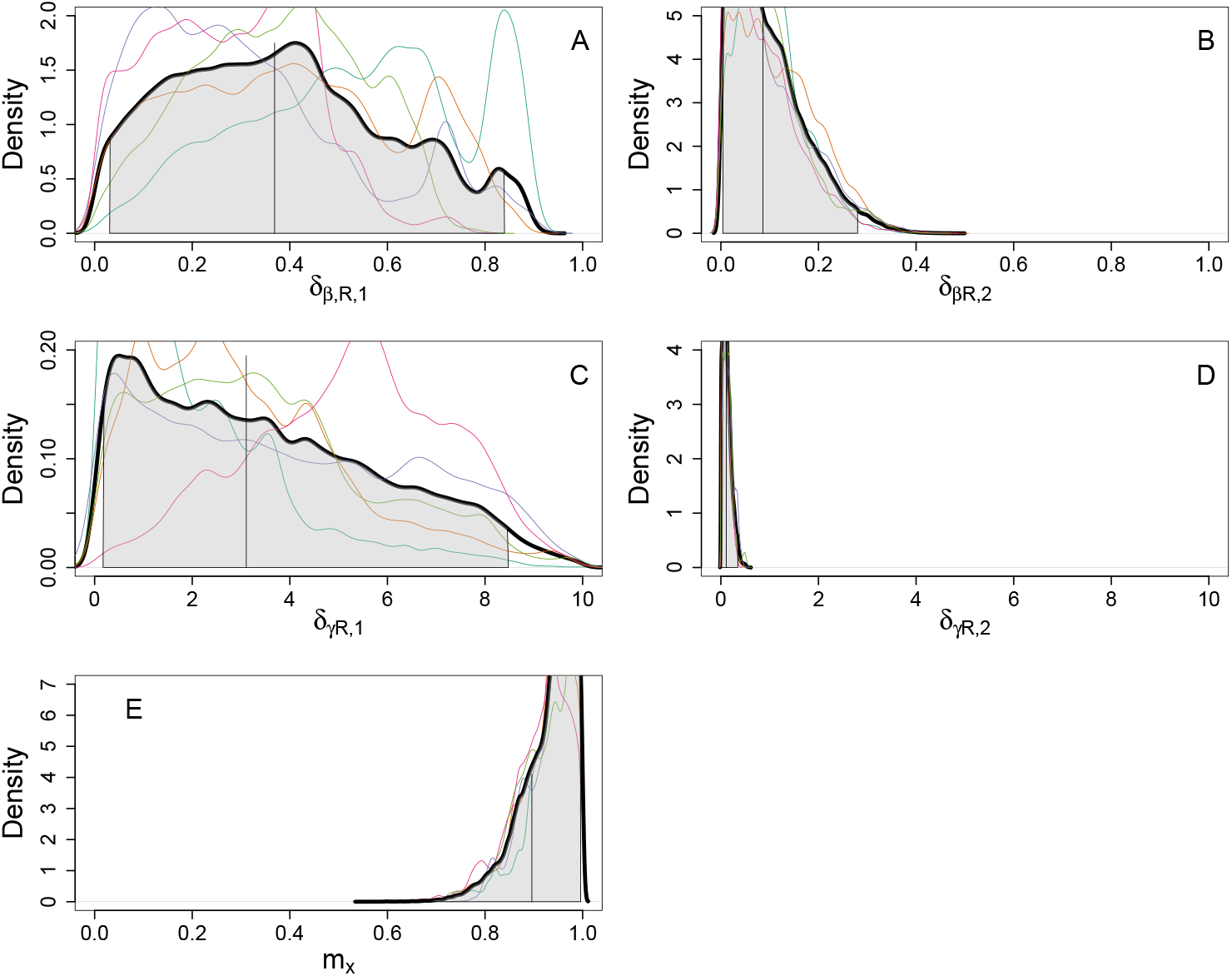
Posterior distributions after inference for the two strain model. Setting and parameters similar to Fig. 3.

**Figure S6:**
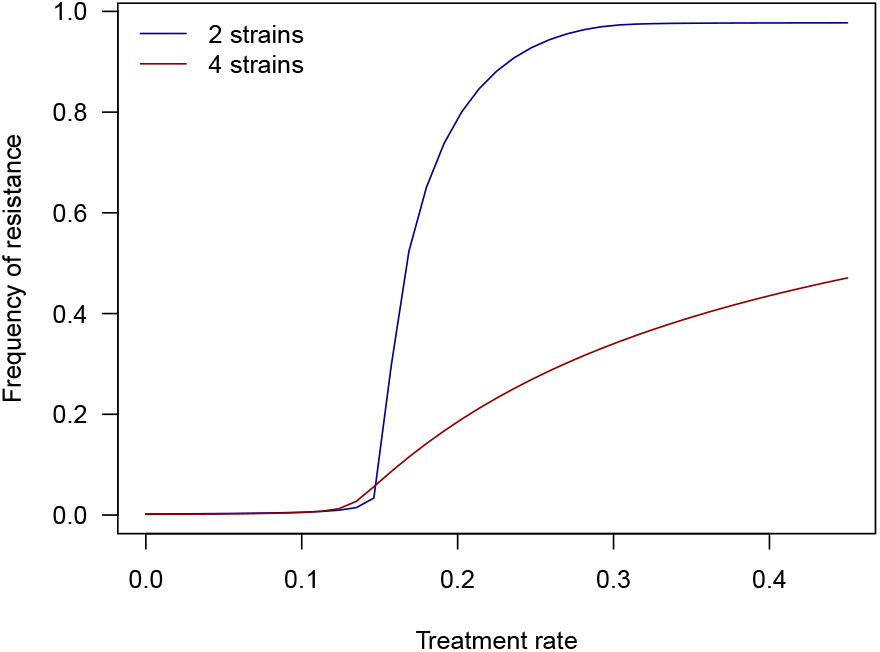
Sensitivity of the frequency of resistance to the treatment rate in the community predicted by the four-strain model (dark red, including genetic structuring as in main text) and the two-strain model (dark blue, without genetic structuring). All other parameters as in Table 1 and mean inferred values from the posterior distributions.

**Figure S7:**
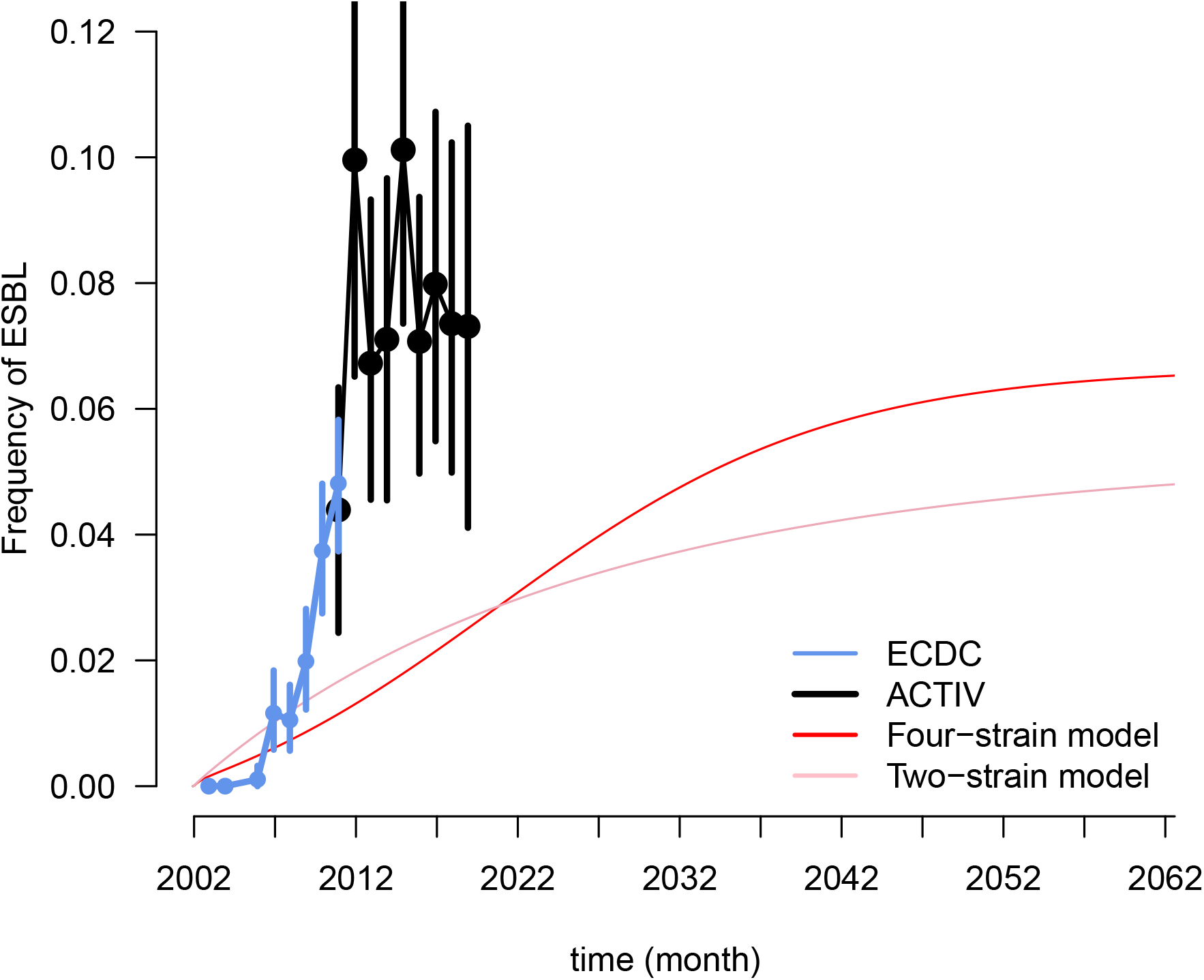
Dynamics of R frequency predicted by the two-strain and four-strain model and comparison with observed dynamics (as in Fig. 2). Red: four-strain model with genetic structuring (main text). Pink: two-strain model (without genetic structuring).

label of figure S7 : time in years

#### Dynamics of R frequency

Expanding the model to include genetic structuring along the persistence-colonization trade-off (Morel-Journel et al., 2025) partially resolved the above issues. Coexistence between sensitive and resistant strains was allowed for a wide range of treatment rates (Fig. S6). Assuming that the R strain was initially brought by travelers in a resistance-free community, the dynamics of R frequency to equilibrium was also faster in the four-strain model (compare red and pink in Fig. S7), though still slower than the observed one (Fig. S7). Adding genetic structuring increased negative-frequency dependence selection on the resistance locus (through linkage with the locus underlying within-host persistence, Fig. S8). Indeed, we found a strong association between the loci determining antibiotic resistance and within-host persistence. Resistance is more beneficial in persistent genotypes, which are more likely to be exposed to antibiotics (Lehtinen et al., 2017). Resistance was strongly associated with persistence, whereas sensitivity was strongly associated with colonization (Fig. S8).

**Figure S8:**
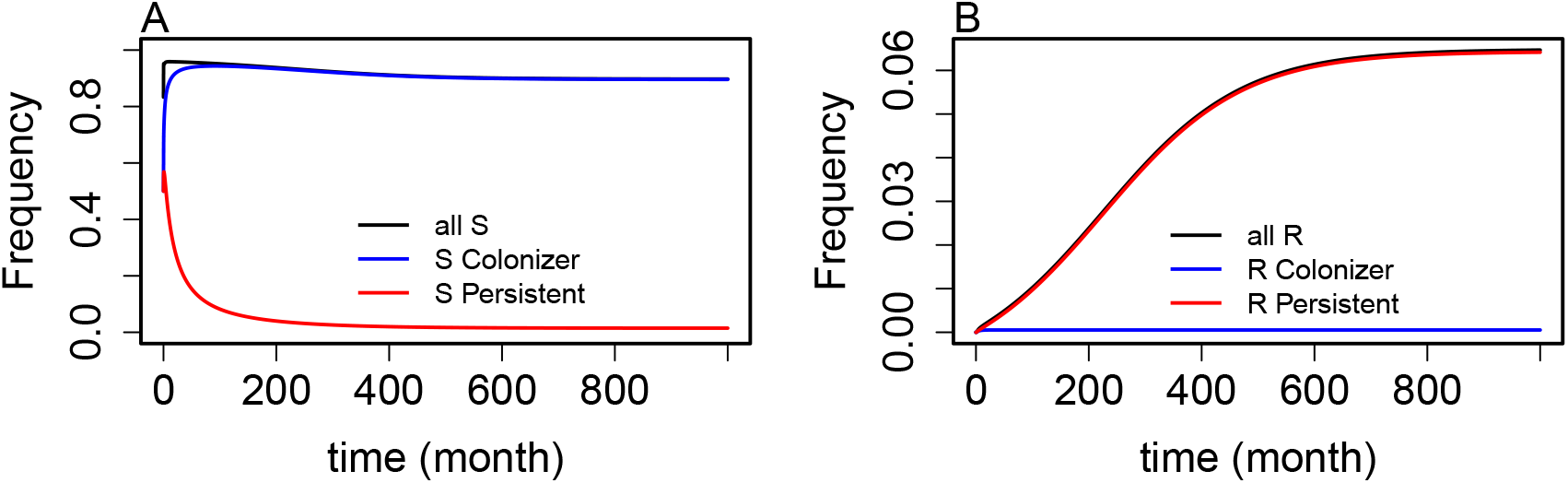
Strain dynamics following the introduction of resistance by travelers at *t* = 0. A: sensitive strains. B: resistant strains. Same parameters as in Table 1.

### S8 Supplementary figures

**Figure S9:**
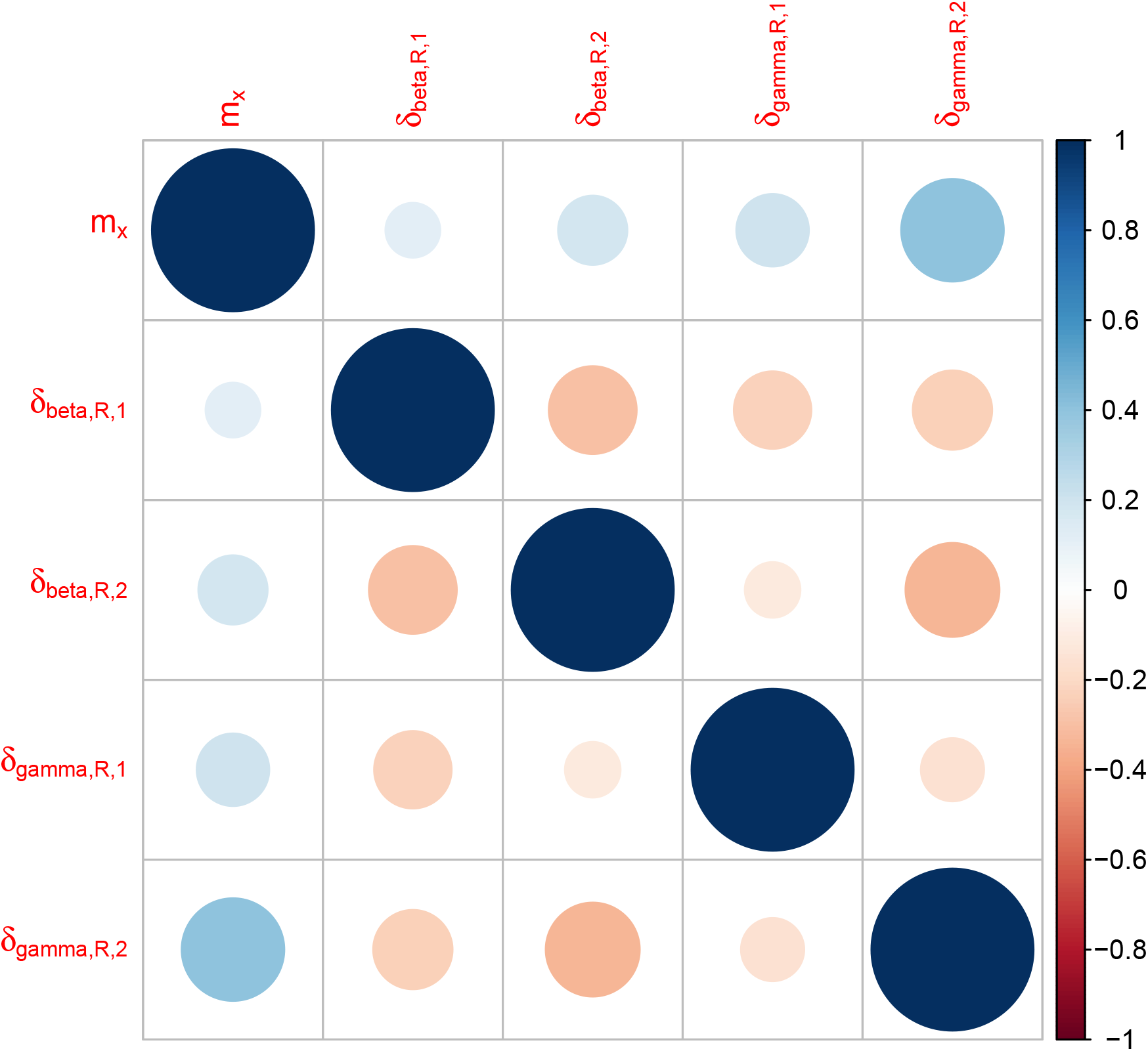
Correlation plot for the parameters in *P* when all MCMC chains are pooled.

**Figure S10:**
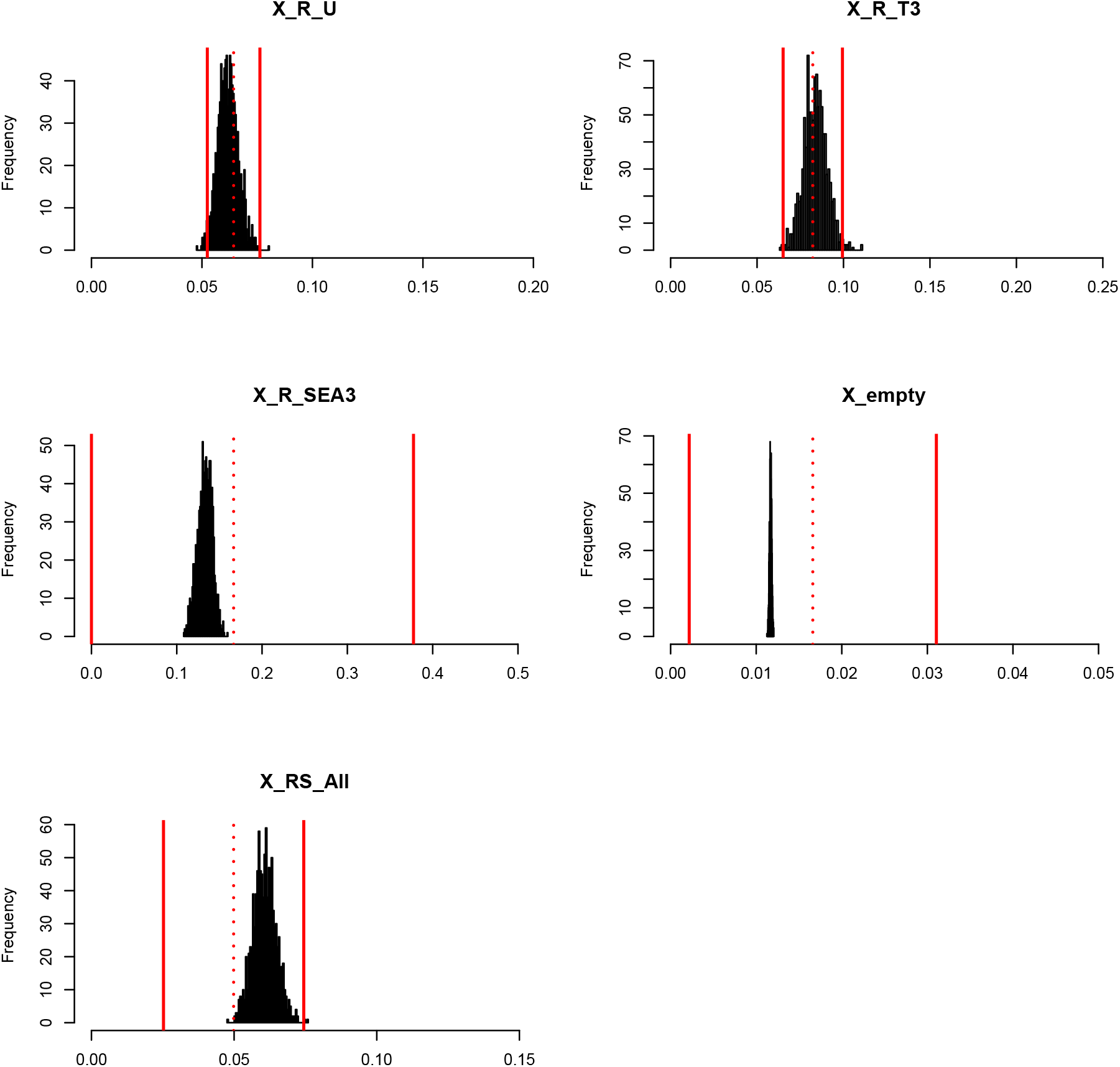
Goodness of fit of the model. Histograms show the distribution of the frequencies of each type of hosts predicted by the model. The dashed-red vertical line show the frequencies of each type of hosts observed in the data set. The solid lines show the 95% CI for the observed frequencies, assuming that the count for each type of hosts is drawn in a binomial distribution B(n,p) with n = number of the corresponding type of host observed and p = the observed frequency of the host type.

**Figure S11:**
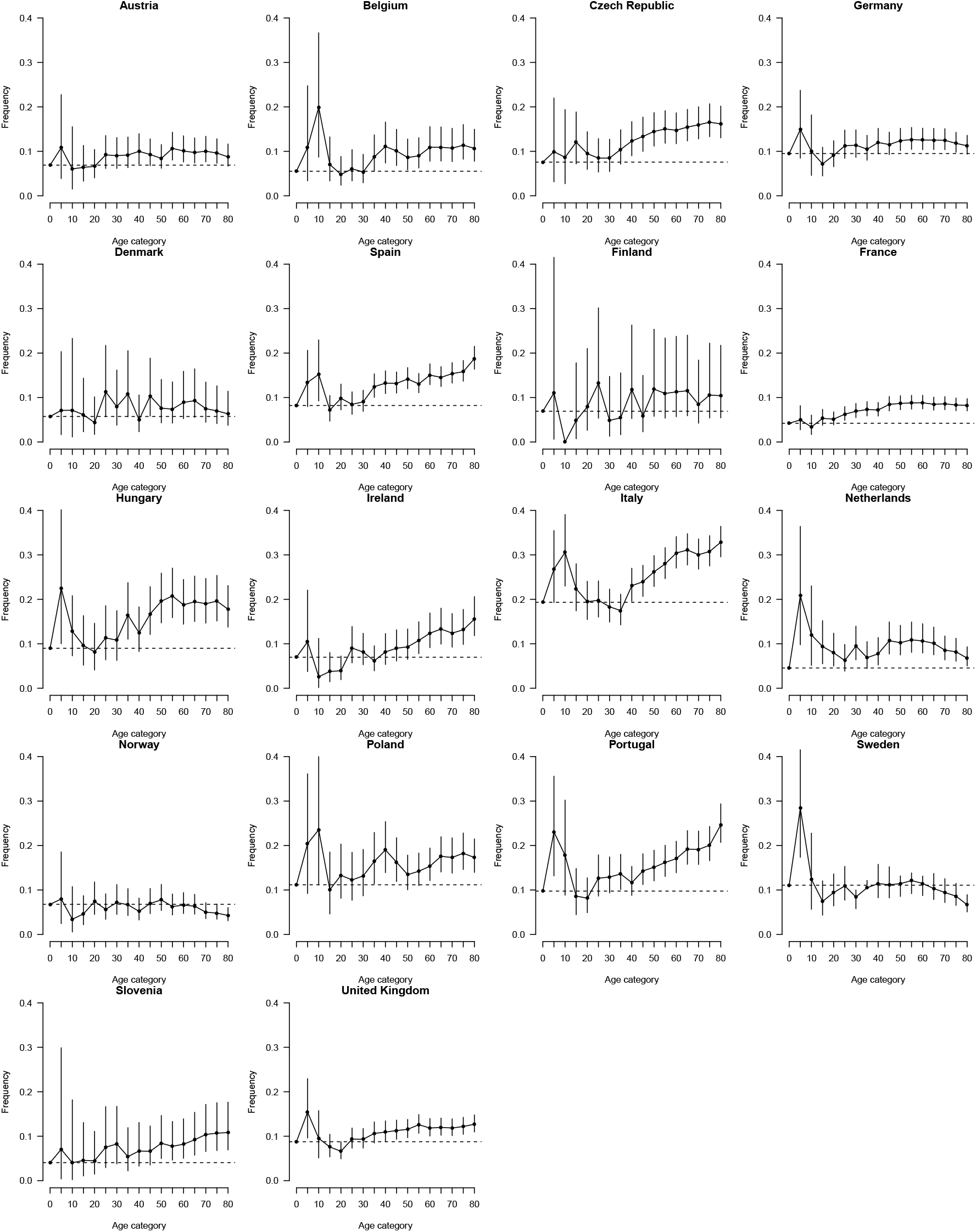
Frequency of ESBL as a function of age in *E. coli* infections in European hospitals. We used data from the ECDC on the frequency of resistance to in *E. coli* isolates in invasive infections (blood, cerebrospinal fluid). We fitted to these data a logistic model for resistance status as a function of year as factor, type of patient (inpatient or outpatient), and age category. We show the predicted resistance for an inpatient infected in 2019, as a function of age category, with 95% confidence intervals. The dashed line is for the [0-4] age category.

## Notes

### Competing Interest Statement

The authors have declared no competing interest.

### Author Declarations

The study was approved by the Saint Germain en Laye Hospital Ethics Committee (10/10/2010-CPP06063). Written informed consent from parents or guardians was obtained.

